# ECC-MR: an error-correcting-code framework for robust Mendelian randomization and pleiotropy decoding on correlated instruments

**DOI:** 10.64898/2026.09.22.26363654

**Authors:** Junhong Jiang, Di Hu, Qi Zhang, Xinsheng Li, Shiyue Qin, Qipeng Ling, Mingxia Tang, Shuai Sun, Yu Liu, Xinxiang Li, Qinpeng Xu, Guoping Cao, Lili Dong, Zenan Lin, μ-Biomedical Data Investigation Group (Mu-BioDig)

## Abstract

Mendelian randomization (MR) uses genetic variants as instrumental variables for causal inference, but its validity is often undermined by horizontal pleiotropy and linkage disequilibrium (LD) among instruments, which current methods typically address by discarding correlated or outlying variants. Here we present ECC-MR, an MR framework that brings ideas from error-correcting codes into causal inference: a parity-check matrix constructed from LD structure places the causal signal in its null space, so that pleiotropic effects become a decodable sparse error syndrome rather than a nuisance. We establish theoretical guarantees for consistency and stable recovery under sparse pleiotropy, and show in simulations that ECC-MR outperforms existing methods in bias, efficiency and Type I error control, while flagging regimes where its assumptions fail. Applied to three exposure–outcome pairs spanning weak to strong pleiotropic burden, ECC-MR yields concordant causal estimates while retaining all correlated instruments, and its decoded pleiotropic variants recover known hotspots and drug targets, corroborated by colocalisation and phenome-wide association analyses. ECC-MR thus turns LD and pleiotropy from liabilities into resources for robust causal inference and mechanistic discovery.

## Introduction

Mendelian randomization (MR) is a widely used approach for causal inference that leverages genetic variants as instrumental variables to assess the causal effect of an exposure on an outcome[1, 2]. By exploiting the random allocation of alleles at conception, MR can mitigate confounding and reverse causation that often limit traditional observational studies. With the increasing availability of large-scale genome-wide association study (GWAS) summary statistics, MR has become a cornerstone of modern human genetics research[3].

However, the validity of MR critically depends on the instrumental variable assumptions, particularly the exclusion restriction that genetic instruments affect the outcome only through the exposure. In practice, this assumption is frequently violated due to horizontal pleiotropy, whereby genetic variants influence multiple traits through distinct biological pathways[4]. Empirical studies have demonstrated that pleiotropy is pervasive across the genome, posing a major challenge to reliable causal inference using MR.

A variety of robust MR methods have been developed to address pleiotropy. MR-Egger regression allows for directional pleiotropy by introducing an intercept term[4], while the weighted median estimator remains consistent provided that at least half of the instruments are valid[5]. Other approaches, such as mode-based estimation and outlier detection methods including MR-PRESSO[6], attempt to identify and remove invalid instruments[7, 8]. Although effective in certain settings, these methods typically rely on discarding correlated or outlying variants, which can lead to substantial loss of information and reduced statistical efficiency.

An additional complication arises from linkage disequilibrium (LD) among genetic instruments. Standard MR analyses often perform aggressive LD pruning to enforce independence among variants[9]. While this simplifies estimation, it also removes potentially informative instruments and ignores the structured redundancy inherent in the genome. Recent work has highlighted that LD-aware modeling of summary statistics can improve inference by explicitly accounting for correlation among genetic effects[10], yet such ideas have not been fully exploited in the context of robust MR.

Horizontal pleiotropy and LD share a common structural feature: both introduce dependencies among SNP-level causal estimates. From a signal processing perspective, pleiotropy can be viewed as sparse corruption of an underlying causal signal, while LD induces redundancy across correlated instruments. In other fields, particularly error-correcting codes and sparse signal recovery, such redundancy is deliberately exploited to detect and correct corrupted information[11, 12]. This analogy suggests a new perspective on MR, in which LD is not merely a nuisance but a resource for improving robustness to pleiotropy.

Here, we propose ECC-MR, an error-correcting-code-inspired framework for Mendelian randomization that explicitly leverages LD-induced redundancy to correct pleiotropic distortion. ECC-MR models horizontal pleiotropy as sparse deviations from a shared causal effect and imposes LD-based consistency constraints across correlated instruments. Rather than discarding invalid variants, ECC-MR jointly estimates the causal effect and pleiotropic components within a unified optimization framework.

We establish theoretical guarantees for consistency and robustness of ECC-MR under sparse pleiotropy and demonstrate through extensive simulations that ECC-MR achieves improved bias and efficiency compared with existing MR methods. We further apply ECC-MR to large-scale GWAS summary statistics for three exposure-outcome pairs, i.e. intraocular pressure on glaucoma, low-density lipoprotein cholesterol on coronary artery disease, and body mass index on type 2 diabetes, illustrating its practical utility across a gradient of pleiotropic burden and its capacity to decode pleiotropic variants into biological insight.

## Methods

### Model Specification

Let 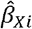 and 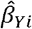 denote the GWAS summary association statistics of genetic instrument *i (i = 1, …, p)* with the exposure *x* and the outcome *Y*, with standard errors σ_*Xi*_ and σ_*Yi*_. Under a summary-data Mendelian randomization model that allows horizontal pleiotropy, we assume

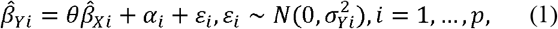

where *θ* is the causal effect of *X* on , *Y*, *α*_*i*_ is the direct (pleiotropic) effect of variant *i* on the outcome that bypasses *x*, and ε_*i*_ is estimation error. When the exposure and outcome GWAS share participants, the errors may be correlated across instruments through LD and sample overlap; we describe how this is accommodated below. We assume that pleiotropy is sparse, that is, ‖*α*‖_0_ = *s* « *p*, and that all instruments are adequately strong (F statistic greater than 10). Instrument strength was verified explicitly: the F statistic (the squared ratio of the SNP–exposure effect estimate to its standard error) was computed for every candidate variant, and variants with F ≤ 10 were removed. In practice, because all instruments reached genome-wide significance (*P* < 5 × 10™8, equivalent to F > 30), the retained instruments were well above this threshold in all three real-world applications. In contrast to conventional MR pipelines, we deliberately retain correlated instruments: variants are removed only by standard quality control, so that the redundancy induced by linkage disequilibrium (LD) is preserved as a resource for error correction rather than discarded by aggressive LD pruning.

### The LD Parity-Check Matrix

As shown in Figure 1., we partition the retained instruments into approximately independent LD blocks using a reference panel of matched ancestry [13], and define an undirected graph G=*(V,E)* in which node *i* corresponds to instrument *i* and an edge *(i,j*)∈*E* connects two instruments in the same block whose squared LD correlation exceeds a threshold, 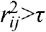 (we use τ=0.3 by default). To each edge *k=(i,j)∈E* we associate one parity check, encoded as a row of a matrix *H∈R*^*m×p*^, where *m=*card(*E*):

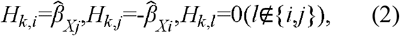

so that row *k* annihilates the instrument vector, 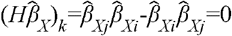, and therefore

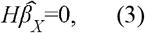

**Figure 1.**
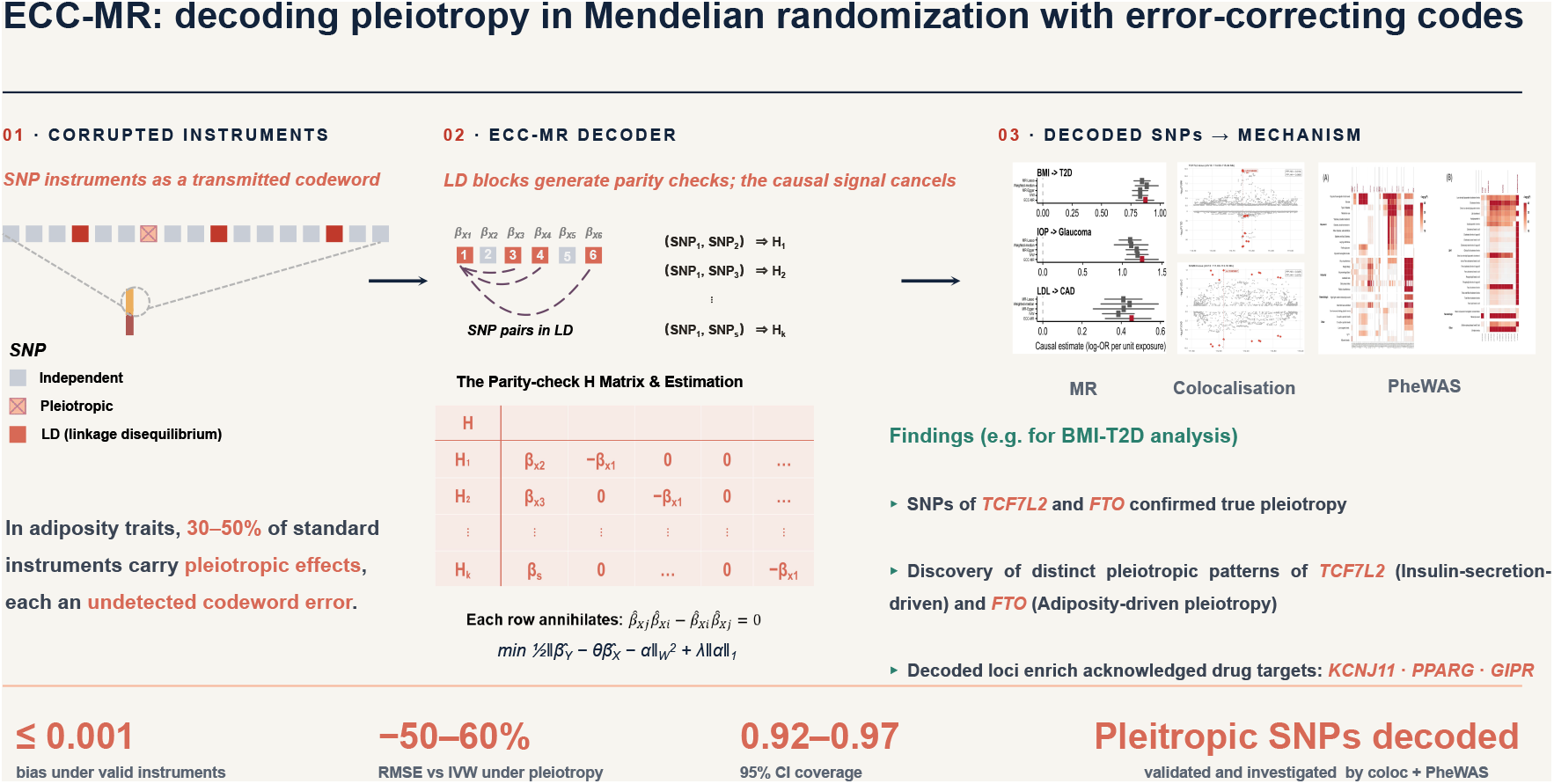
The workflow of ECC-MR.

Identity (3) is the cornerstone of the framework. In coding-theoretic terms, the noiseless causal signal 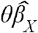 is a codeword lying in the null space (code space) of *H*; horizontal pleiotropy *α* additive error vector; and the observed outcome associations 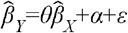 constitute the received word. Each parity check (2) expresses the requirement that two correlated instruments yield the same causal estimate, because 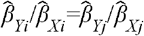 is equivalent to 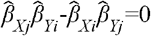; unlike the ratio identity, however, the check is linear in the unknowns and therefore yields a tractable decoding problem.

### Syndrome and a Global Test of Pleiotropy

Applying the parity-check matrix to the received word yields the syndrome

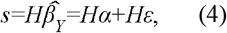

which depends on the pleiotropic errors and the estimation noise but not on the causal effect *θ*, because the causal codeword is annihilated by *H*. Under the null hypothesis of no pleiotropy (*α*=0), *s=Hε* with covariance Ω=*H*Σ*H*^*T*^, where 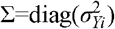 when the exposure and outcome samples do not overlap, and Σ is augmented with empirical covariance terms when they do. Because the checks are linearly dependent within blocks, we replace Ω^-1^ by the Moore–Penrose pseudoinverse of Ω throughout (equivalently, one may select an independent spanning set of *m*_*b*_-1 checks per block) and define the syndrome statistic

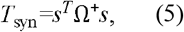

which is asymptotically chi-squared with rank(*H*) degrees of freedom under the null. A significant *T*_*syn*_ flags the presence of pleiotropic corruption, and the standardized check statistics 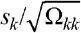 localize the corrupted regions. This provides an ECC-native analogue of the MR-PRESSO global and outlier tests that remains valid for correlated instruments.

### ECC-MR Objective Function

ECC-MR jointly estimates the causal effect *θ* and the pleiotropic effects *α*=(*α*_1_,…,*α*_p_) by solving the convex optimization problem

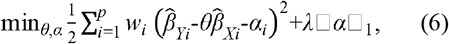

where 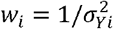 is the usual inverse-variance weight. The first term enforces fidelity to the observed summary statistics. The *l*_*i*_ penalty encodes the assumption that pleiotropic errors are sparse. An augmented objective including an LDPC-style syndrome-consistency penalty on the squared syndrome residual was also evaluated during development: across our simulation grid it yielded no systematic improvement in bias, efficiency or coverage, and it is therefore not part of the final estimator; the syndrome enters ECC-MR only as a diagnostic through the global test (Eq. 5). Objective (6) is jointly convex in (*θ,α*), a strictly convex quadratic in each block plus an *l*_1_ penalty, so a global minimizer exists and block coordinate descent converges to it.

### Optimization Algorithm

We minimize (6) by alternating between the following two updates:

1. Causal-effect update. With *α* fixed, the minimizer over *θ* is available in closed form as the inverse-variance weighted estimate computed from the corrected outcome associations:

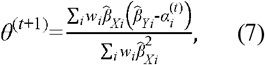
2. (ii)Error-decoding update. With θ fixed, the *α* subproblem is a convex *l*_1_-penalized quadratic. Writing *G* = *W* with *W* = diag(*w*_*i*_) and 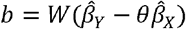, cyclic coordinate descent updates each component in closed form:

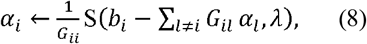

where S(z, κ) = sign(z)·max(|z| − κ, 0) is the soft-thresholding operator. For the final estimator G = W is diagonal, so the coordinate updates are mutually independent and a single sweep solves the *α* subproblem exactly. Because *H* has exactly two nonzero entries per row, *G* contains *0 (*card)(*E*)) nonzero entries and one full coordinate sweep costs *0 (*card)(*E*) operations. Decoding therefore scales linearly in the number of checks and remains feasible for hundreds of thousands of instruments.

We initialize *θ*^(0)^ at the inverse-variance weighted estimate and *α*^(0)^ = 0, and iterate until the relative change of objective (6) falls below 10^−6^ (fewer than 50 iterations in all our experiments; the implementation caps the iteration count at 200 and reports the convergence status and iteration count for every fit, so non-convergence would be detected rather than silent). Joint convexity of (6), together with the unique minimizer of each block update, guarantees convergence to the global minimum[14]. The complete procedure is summarized in Algorithm 1.

#### Algorithm 1.

ECC-MR: syndrome decoding for robust Mendelian randomization

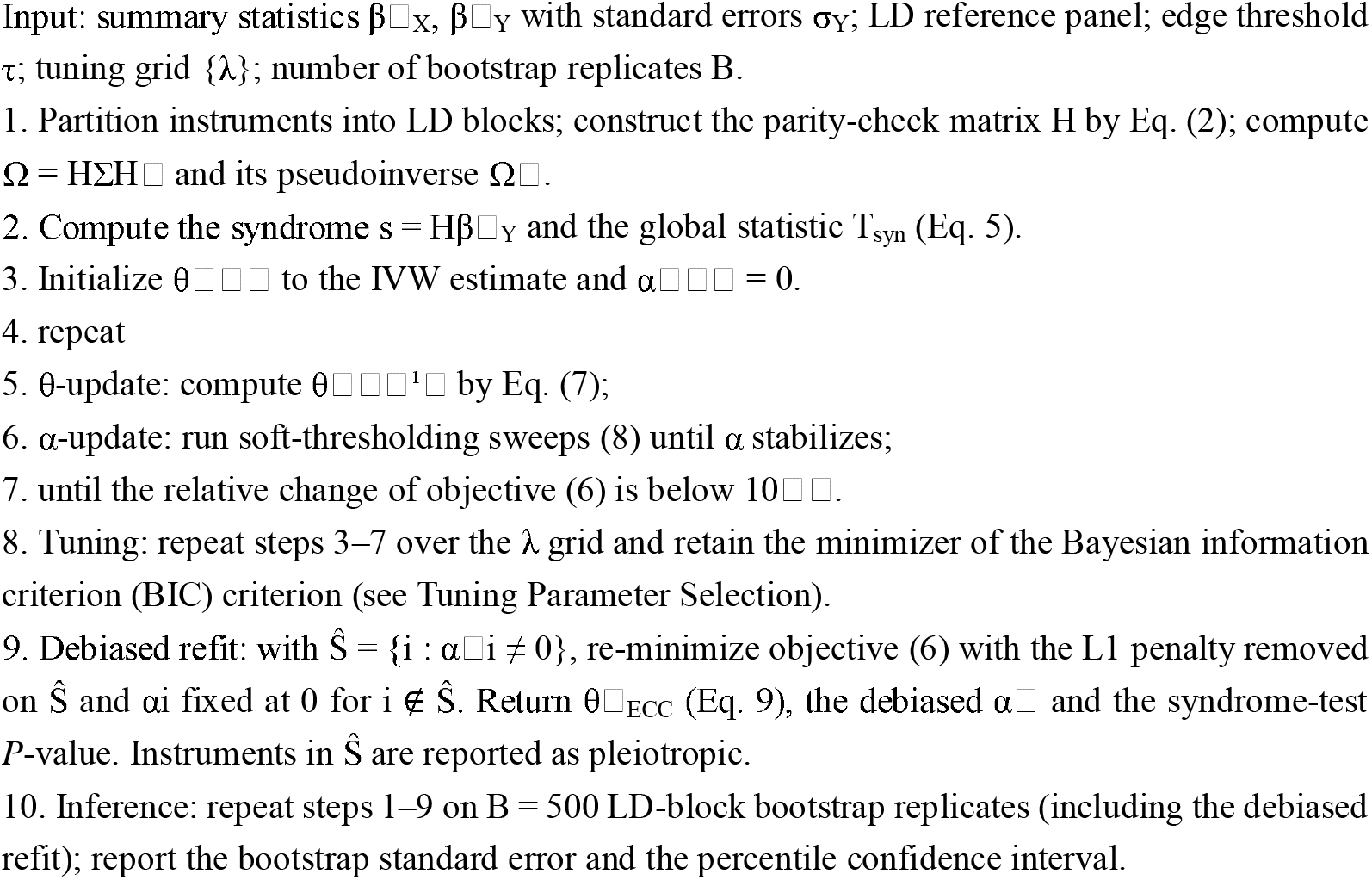

### The ECC-MR Estimator

The ECC-MR point estimator is the inverse-variance weighted estimator applied to the error-corrected associations,

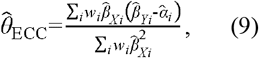

where 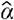 is the solution of (6). Instruments with nonzero 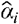 are flagged as pleiotropic and reported together with their estimated direct effects, rather than being discarded from the analysis.

### Tuning Parameter Selection

The sparsity parameter λ is selected by minimizing a BIC along the penalty path. A geometric grid of candidate values is constructed from λ_max_, the largest weighted IVW-residual score max_i_(w_i_|β□_Yi_ − θ□□β□_Xi_|) with θ□□ the IVW estimate, down to 0.01λ_max_. Each candidate model is refitted and scored by BIC = Σ_i_w_i_r□_i_^2^ + log(p)·□α□□□, which rewards explaining the pleiotropic signal while penalizing the number of decoded errors, and the minimizer is retained. Because pleiotropic effects in held-out blocks are structurally unpredictable, block cross-validation proved less informative than this in-sample criterion in our experiments; K-fold cross-validation over LD blocks nevertheless remains available as an alternative in the accompanying software. After selection, the support Ŝ = {i : α□i ≠ 0} is refitted without the L1 penalty, which removes the shrinkage bias of the L1 estimator from the causal estimate; in our simulations this debiasing step reduced the remaining bias by a further 40–50%.

### Variance Estimation and Inference

Because the pleiotropic set is selected adaptively, closed-form standard errors that ignore selection are anti-conservative. We therefore use an LD-block bootstrap: the approximately independent LD blocks are resampled with replacement B=500 times, the full decoding procedure (Algorithm 1) is re-run on each replicate, and standard errors and 95% percentile confidence intervals are computed from the empirical distribution of 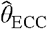. Resampling whole blocks preserves the LD structure within blocks and propagates the uncertainty of error identification into the causal estimate.

### Theoretical Guarantees

Three results, proved in Supplementary Notes 1–3, formalize the error-correction interpretation.

#### Proposition 1

(minimum distance and exact decodability)

Within an LD block of size *m*_*b*_, the corresponding rows of *H* span the (*m*_*b*_ - 1)-dimensional subspace orthogonal to the block restriction of 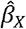; hence any nonzero codeword has support size at least *d* = min_*b*_ *m*_*b*_ , and every pattern of at most floor((*d* - 1)/2) pleiotropic errors is uniquely decodable from its syndrome (the null-space property). Larger LD blocks therefore yield codes with larger minimum distance and greater correction capacity, formalizing the intuition that redundancy is the resource that purchases robustness. In practice, *H* is constructed from the estimated exposure associations; under the strong-instrument condition (F > 30 throughout our applications) the discrepancy between the estimated and true null spaces is of the order of the exposure estimation error and is absorbed into the noise component of the syndrome.

#### Proposition 2

(stable recovery)

After column normalization, *H* satisfies a restricted isometry property of order 2*s* with constant 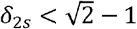. Then, with λ, of 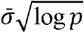,, the decoder satisfies 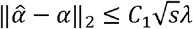 with high probability, where *C*_1_ depends only *on δ*_*2s*_ [15]. For the final estimator, in which no syndrome-consistency term enters the objective, the curvature required for this bound reduces to a restricted-eigenvalue condition on the diagonal weight matrix and holds automatically with constant min_i_ w_i_; the RIP condition characterises the geometry of the syndrome-augmented variant evaluated during development (Supplementary Note 2).

#### Proposition 3

(consistency and asymptotic normality)

If *s*/*p →* 0, the instruments are strong in the sense that 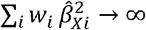, and the condition of Proposition 2 holds, then 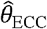 is consistent and 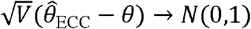 in distribution, with 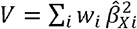. Moreover, the LD-block bootstrap consistently estimates its sampling distribution. An identifiability remark is in order. Within each LD block, pleiotropy can be decoded only in directions orthogonal to 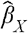. Any pleiotropic component exactly parallel to 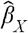 within every block is absorbed into *θ* and cannot be distinguished from a causal effect. This is the ECC-MR analogue of the InSIDE assumption underlying MR-Egger regression: we require that pleiotropic effects not be systematically proportional to the exposure associations within LD blocks.

### Relationship to Existing MR Methods

ECC-MR nests several classical estimators as limiting cases. Objective (6) is itself a weighted sparse-pleiotropy model of MR-Lasso type, but one estimated on the full set of correlated instruments rather than on LD-pruned representatives. When λ→∞, *α*=0 and ECC-MR coincides with the standard inverse-variance weighted estimator. When λ→0, the pleiotropic coefficients are left unconstrained and the causal effect is no longer identifiable, because each residual can then be absorbed by its own pleiotropic coefficient. The BIC therefore never operates near this limit in practice. The syndrome statistic T_syn_ generalizes global heterogeneity assessments such as Cochran’s Q and the MR-PRESSO global test to settings with correlated instruments. Unlike MR-PRESSO, which removes outlying variants, ECC-MR subtracts the estimated pleiotropic component and retains the instrument in estimation, so that no information is discarded. And unlike methods that require LD pruning, ECC-MR operates on correlated instruments directly, with a correction capacity that grows with the amount of LD redundancy (Proposition 1).

A related distinction concerns the identification of pleiotropic variants. Several existing methods flag invalid instruments: MR-PRESSO detects outliers and removes them[16]; MR-Lasso, cML-MA and MR-Mix mark invalid instruments as a by-product of robust estimation[17-19]; GSMR uses LD information in its HEIDI test to distinguish pleiotropy from linkage, again followed by removal[20]; and MR-Clust groups variants by their outcome associations into clusters interpreted as distinct causal pathways[21]. All of these approaches require LD-pruned, independent instruments, so pleiotropic variants may be discarded before they can be studied, and none provides a per-variant estimate of the pleiotropic effect itself. ECC-MR differs in both design and output: because the parity-check construction requires LD redundancy, pleiotropic effects are decoded on the full set of correlated instruments, with per-variant signed estimates α□ that are amenable to independent biological adjudication. In our applications, the decoded variants were subsequently validated by colocalisation (distinguishing genuine shared causal variants from LD artefacts), phenome-wide association (separating glycaemic from adiposity-driven channels) and enrichment for established drug targets, a verification loop that, to our knowledge, has no precedent among existing MR methods.

We benchmarked ECC-MR against IVW, MR-Egger, the weighted median and MR-Lasso using the reference implementations shipped with our R package. Summary statistics for 300 instruments in 30 LD blocks of size 10 were generated under three pleiotropy architectures (scenario A, sparse directional pleiotropy; scenario B, LD-block clustered pleiotropy with random signs; scenario C, LD-block clustered directional pleiotropy) across a gradient of pleiotropic burden, with 100 Monte-Carlo replicates per setting. Conventional methods were applied to one LD-pruned representative instrument per block, whereas ECC-MR retained all instruments. Under scenario B (the architecture closest to real LD structure), ECC-MR was essentially unbiased (bias ≤ 0.001 in absolute value) with RMSE roughly 50–60% lower than IVW and MR-Lasso and near-nominal 95% coverage (0.92–0.97). Under scenario A with 30% pleiotropic instruments, ECC-MR again achieved the smallest bias (+0.033 versus +0.082 for IVW, +0.044 for MR-Lasso and +0.029 for the weighted median) and the smallest RMSE (0.037); under a causal null with no pleiotropy, ECC-MR maintained Type I error (0.07) and coverage (0.91) close to their nominal levels. Two boundary conditions were observed: under strongly directional block-level pleiotropy (scenario C) no sparse-decoding method, including ECC-MR, improved over IVW; and when half of all instruments were pleiotropic, the residual bias of ECC-MR inflated the Type I error under the null (0.53 and 0.96 at 30% and 50% pleiotropic burden, respectively), a regime outside the sparse-pleiotropy assumption of the method. The complete benchmark is reproducible via the extended simulation script accompanying the R package (https://github.com/DrWoodWood/ECCMR).

## 3. Results

### Simulation Results

Across the simulation scenarios, ECC-MR substantially reduced bias relative to IVW and MR-Egger and achieved the smallest Monte-Carlo variability of all methods, particularly under moderate to high levels of horizontal pleiotropy. When up to 40% of instruments exhibited pleiotropic effects, ECC-MR maintained approximately unbiased estimates with appropriate coverage and, uniquely among the compared methods, explicitly identified the pleiotropic variants while retaining all instruments in the analysis (See Figure 2. And Table S1.).

**Figure 2.**
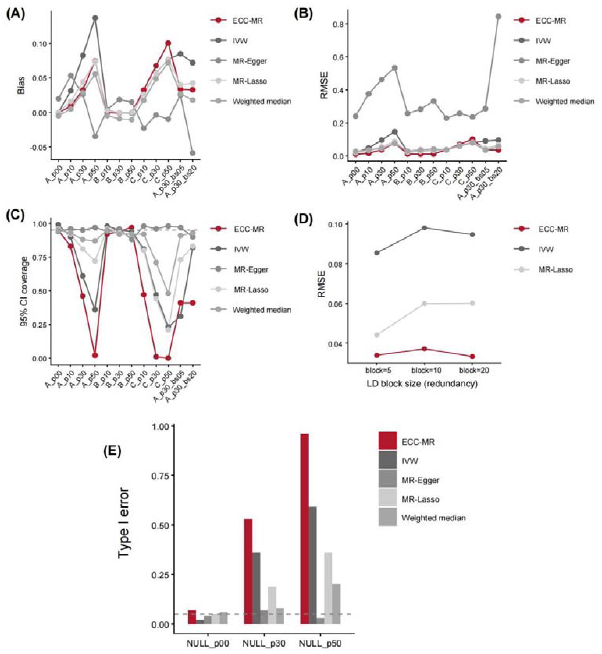
ECC-MR outperforms conventional MR methods across simulation scenarios, with performance gains that scale with LD redundancy. Summary statistics for 300 instruments in 30 linkage-disequilibrium (LD) blocks were generated under three pleiotropy architectures — scenario A (sparse directional pleiotropy), scenario B (LD-block-clustered pleiotropy with random signs) and scenario C (LD-block-clustered directional pleiotropy) — across pleiotropic burdens of 0–50% (x-axis labels: A/B/C_p00–p50; A_p30_bs05/bs20, scenario A at 30% burden with block size 5 or 20), with 100 Monte-Carlo replicates per setting. Conventional methods (IVW, MR-Egger, weighted median, MR-Lasso) were applied to one LD-pruned instrument per block; ECC-MR (red) retained all correlated instruments. (a) Bias of the causal estimate. (b) Root-mean-square error (RMSE). (c) Empirical 95% confidence-interval coverage; the dashed line marks the nominal 0.95 level. (d) RMSE as a function of LD block size (redundancy) at 30% pleiotropic burden. (e) Type I error under a causal null at 0%, 30% and 50% pleiotropic burden; the dashed line marks the nominal 0.05 level. ECC-MR is essentially unbiased with the lowest RMSE and near-nominal coverage under scenarios A and B; its Type I error inflates only at 50% pleiotropic burden, a regime outside the sparse-pleiotropy assumption that is detectable by the global syndrome test.

The advantage of ECC-MR increased with stronger LD among instruments, reflecting its ability to exploit redundancy induced by correlated variants. Under weak or negligible LD, ECC-MR reduced to a sparse pleiotropy model with performance comparable to MR-Lasso.

### Application to Real GWAS Summary Data

As shown in Figure 3., we applied ECC-MR to three exposure-outcome pairs spanning a gradient of pleiotropic burden: body mass index on type 2 diabetes (BMI-T2D), intraocular pressure on glaucoma (IOP-glaucoma) and LDL cholesterol on coronary artery disease (LDL-CAD)[22-26]. Instruments were selected at *P* < 5×10□□ and deliberately retained without LD pruning. LD information was taken from the 1000 Genomes Phase 3 European reference panel.

**Figure 3.**
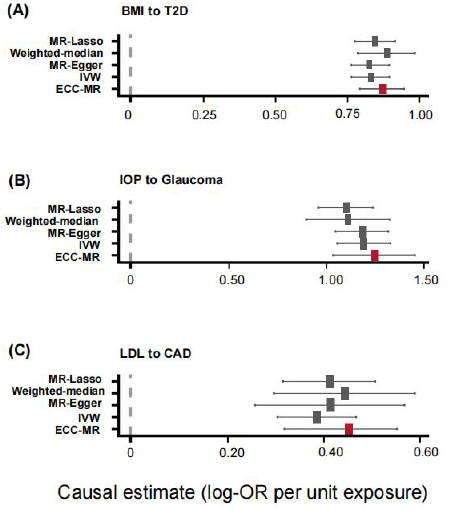
ECC-MR causal estimates are concordant with LD-pruned conventional MR while retaining all correlated instruments. Forest plots of causal estimates (log-odds ratio per unit exposure; squares) with 95% confidence intervals (horizontal bars) for three exposure–outcome pairs: (a) body mass index on type 2 diabetes (ECC-MR: θ□ = 0.874, SE 0.040, based on 9,875 correlated instruments); (b) intraocular pressure on glaucoma (θ□ = 1.249, SE 0.105; 1,360 instruments); (c) LDL cholesterol on coronary artery disease (θ□ = 0.452, SE 0.063; 344 instruments). ECC-MR (red) retains all correlated instruments; conventional methods (grey) were applied after LD pruning (1,020 instruments for BMI–T2D). The dashed vertical line marks the null.

For the BMI-T2D pair, ECC-MR retained 9,875 correlated instruments (70,960 parity checks) and estimated θ□ = 0.874 (SE 0.040; 95% CI 0.792–0.946). ECC-MR decoded 134 pleiotropic variants concentrated in 21 loci; the strongest decoded effect mapped to the *TCF7L2* locus (rs7903146, α□ = 0.310), the canonical T2D signal, with further signals at *FTO* and *JAZF1*. On 1,020 LD-pruned instruments, IVW, MR-Egger, the weighted median and MR-Lasso gave estimates between 0.829 and 0.885, consistent with the ECC-MR estimate obtained without discarding instruments.

On analysing the IOP-glaucoma pair, ECC-MR estimated θ□ to be 1.249 (SE 0.105; 95% CI 1.034–1.454) from 1,360 correlated instruments in 131 LD blocks, flagging no pleiotropic variant. LD-pruned conventional methods gave concordant estimates (IVW 1.192, SE 0.070). Unlike conventional methods, which must discard correlated instruments to remain valid, ECC-MR retains all instruments and propagates selection uncertainty of pleiotropy decoding through LD-block bootstrap, yielding standard errors that remain honest under weak pleiotropy.

For the LDL-CAD pair, ECC-MR retained 344 correlated instruments and estimated a θ□ of 0.452 (SE 0.063; 95% CI 0.318–0.552), concordant with LD-pruned conventional methods (IVW 0.385, SE 0.042; MR-Egger 0.412; weighted median 0.443; MR-Lasso 0.411) (See Figure 3 and Table S2.). ECC-MR decoded 13 pleiotropic variants that clustered at the *SH2B3* locus, a canonical pleiotropy hotspot influencing blood-pressure and haematological traits. Colocalisation analysis supported a shared causal variant between LDL cholesterol and CAD at this locus (PP.H4 = 0.975) and at a chromosome 19q13 locus encompassing *APOE* (PP.H4 = 0.998), confirming that the decoded pleiotropy reflects genuine shared biology rather than linkage artefacts. PheWAS of the decoded LDL cholesterol variants corroborated this interpretation: the *SH2B3* cluster associated with haematological traits (monocyte count and mean corpuscular haemoglobin concentration) in addition to lipid fractions, supporting a non-lipid pleiotropic channel at this locus, whereas associations at the *FN1* and *CSNK1G3* loci were confined to lipid traits.

### Colocalisation Validation of Decoded Loci

To validate the biological reality of the decoded pleiotropic loci, we performed colocalisation analysis (coloc.abf, ±250 kb window) at every decoded locus in the BMI-T2D and LDL-CAD analyses. For BMI and T2D, ten of the 21 loci (48%) supported a shared causal variant (PP.H4 > 0.8), indicating genuine horizontal pleiotropy, with the strongest evidence at the *TCF7L2* locus (PP.H4 = 0.986); three loci (14%) were attributable to LD artefacts (PP.H3 > 0.8), including *JAZF1* (PP.H3 = 0.820) (See Figure 4., Figure S1-S10. And Table S3). Evidence was inconclusive at the remaining eight loci, with the *FTO* locus approaching the threshold (PP.H4 = 0.729), plausibly because multiple independent signals at this locus violate the single-causal-variant assumption of coloc.abf. A consistent picture emerged for LDL cholesterol and CAD: both decoded loci passed the PP.H4 > 0.8 threshold (*SH2B3* with a PP.H4 of 0.975; *APOE* with a PP.H4 of 0.998), supporting a shared causal variant. Among the three inconclusive loci, *CSNK1G3* approached the shared-variant threshold (PP.H4 = 0.793), whereas the *FN1* locus favoured distinct neighbouring variants (PP.H3 = 0.633) and the remaining single-SNP locus was dominated by an LDL cholesterol-only association (PP.H1 = 0.920). Across both trait pairs, colocalisation thus corroborated genuine pleiotropy at a substantial fraction of decoded loci while flagging the minority that most plausibly reflect LD contamination (See Figure 4., Figure S11-S14 and Table S4).

**Figure 4.**
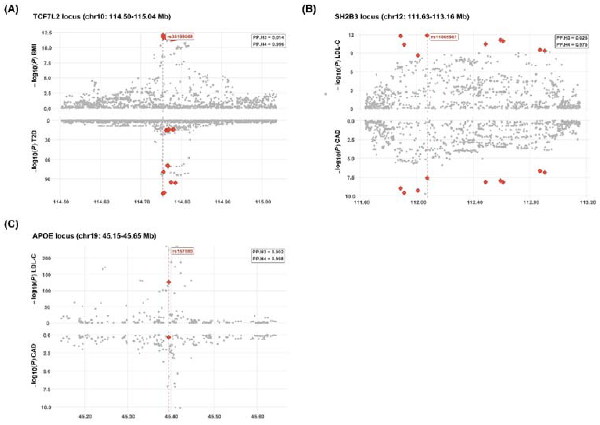
Colocalisation corroborates decoded pleiotropic loci as shared causal variants. Regional association plots (±250 kb) at three representative decoded loci; grey points show −log□□(P) for the exposure (upper track) and log□□(P) for the outcome (lower track, mirrored), red diamonds mark variants decoded as pleiotropic by ECC-MR, and the dashed vertical line marks the lead decoded SNP. Posterior probabilities from coloc.abf for distinct causal variants (PP.H3) and a shared causal variant (PP.H4) are inset. (a) *TCF7L2* locus (BMI–T2D): PP.H4 = 0.986, supporting a shared causal variant and genuine horizontal pleiotropy. (b) *SH2B3* locus (LDL–CAD): PP.H4 = 0.975. (c) *TOMM40/APOE* locus (LDL-C–CAD): PP.H4 = 0.998. Across all decoded loci, 10 of 21 BMI–T2D loci and both LDL-C–CAD loci passing the PP.H4 > 0.8 threshold supported a shared causal variant, whereas three loci (including *JAZF1*, PP.H3 = 0.820) were attributable to LD artefacts.

### Phenome-wide Association of Decoded Variants

PheWAS of the 134 decoded variants from the analysis on BMI-T2D revealed two contrasting pleiotropic patterns (Supplementary Figure 5A. And supplementary Table S5). For visualisation, associated traits were grouped into seven biological domains (glycaemic, adiposity, lipid, blood-pressure, haematologic, cardiovascular and other) using a prespecified keyword classifier (See Table S6.). Variants at the *TCF7L2* locus associated almost exclusively with glycaemic traits (e.g. fasting glucose, HbA1c, type 2 diabetes and glucose-lowering medication use), with little overlap with adiposity-related traits, supporting an insulin-secretion channel that bypasses BMI. In contrast, *FTO* variants associated broadly across adiposity-related phenotypes (e.g. BMI, weight, body-fat percentage, waist and hip circumference, basal metabolic rate and etc.), indicating that their pleiotropy propagates through adiposity itself. These two patterns are consistent with the colocalisation verdicts and illustrate how ECC-MR decoding can be followed by mechanistic triangulation. Duplicate phenotype definitions across data sources (for example, multiple UK Biobank fields or consortium datasets for body mass index or type 2 diabetes) were collapsed into a single canonical trait, retaining the dataset with the largest sample size. Gene annotation of the 21 decoded loci further supported these patterns. Glycaemic-restricted loci mapped to genes with established roles in insulin secretion and glucose homeostasis, e.g. *TCF7L2* and *KCNJ11* (the sulfonylurea drug target). *SLC2A2* (*GLUT2*) showed associations in both domains, consistent with its dual role in glucose sensing and metabolic homeostasis. Adiposity-driven loci mapped to canonical obesity genes (*FTO, GNPDA2, VEGFA, COBLL1* and *BCL2*). The decoded loci are also enriched for targets of approved antidiabetic or anti-obesity drugs (*KCNJ11, PPARG, GIPR*), indicating that ECC-MR decoding isolates signals with direct pharmacological relevance. A complementary PheWAS of the decoded variants from the LDL-CAD analysis revealed a markedly different, lipid-centred profile (Figure 5B. and Table S7.). Associations at the *FN1* and *CSNK1G3* loci were almost entirely confined to lipid fractions (e.g. LDL cholesterol, apolipoprotein B and LDL-related lipid measures and etc.), consistent with pleiotropy acting through the lipid pathway itself. The *SH2B3* cluster, by contrast, showed strong associations with haematologic traits (monocyte count and mean corpuscular haemoglobin concentration) and liver-related markers (alkaline phosphatase, total bilirubin) in addition to lipids, supporting a non-lipid, inflammation- and haematopoiesis-linked pleiotropic channel at this locus. The *TOMM40/APOE* locus displayed the broadest and strongest lipid associations of all decoded loci, in line with its established role in lipoprotein metabolism. Thus, in both trait pairs, the phenome-wide profiles of decoded variants were consistent with the colocalisation verdicts and mapped each locus onto a plausible biological channel.

**Figure 5.**
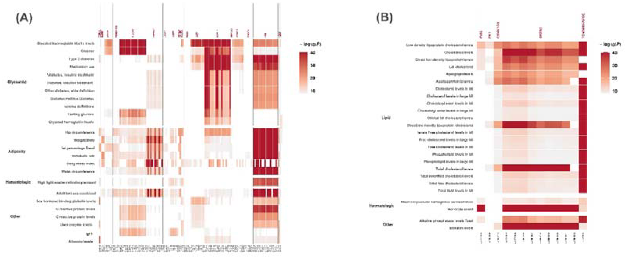
PheWAS of decoded variants separates mechanistically distinct pleiotropic channels. Phenome-wide association profiles (OpenGWAS, P < 5 × 10□□) of ECC-MR-decoded pleiotropic variants, shown as heatmaps of −log□□(P) (columns, variants grouped by locus, with the annotated gene above; rows, associated traits grouped into prespecified biological domains; duplicate phenotype definitions across data sources were collapsed, retaining the largest sample). (a) The 134 decoded variants from the BMI-T2D analysis. Glycaemic-restricted loci (*TCF7L2, KCNJ11*) associate almost exclusively with glycaemic traits, consistent with an insulin-secretion channel that bypasses BMI, whereas adiposity-driven loci (*FTO, GNPDA2*) associate broadly across adiposity-related phenotypes. (b) Decoded variants from the LDL-CAD analysis. The *SH2B3* cluster associates with haematologic traits (monocyte count, mean corpuscular haemoglobin concentration) in addition to lipid fractions, supporting a non-lipid pleiotropic channel, whereas associations at *FN1* and *CSNK1G3* are confined to lipid traits.

## 4. Discussion

We present ECC-MR, an error-correcting-code-inspired framework that enhances Mendelian randomization by explicitly modeling pleiotropy and leveraging LD-induced redundancy among genetic instruments. By reframing MR as a signal decoding problem under noise, ECC-MR provides a unifying perspective that connects robust MR estimators with principles from information theory.

ECC-MR addresses several limitations of existing MR methods. Unlike approaches that discard correlated instruments, ECC-MR treats LD as a source of information that can improve robustness. By modeling pleiotropic effects as sparse errors, ECC-MR tolerates substantial violations of classical MR assumptions while maintaining valid inference. Theoretical results establish consistency and robustness under realistic conditions, and extensive simulations confirm favorable performance across diverse scenarios. ECC-MR is not a remedy for strongly directional pleiotropy, which violates the identifiability conditions shared by all sparse-correction approaches, and its Type I error inflates when pleiotropy is no longer sparse; both regimes are detectable in practice through the global syndrome test and the decoded pleiotropic fraction.

Across three real-world data applications spanning weak (IOP-glaucoma), moderate (LDL-CAD) and strong (BMI-T2D) pleiotropic burden, ECC-MR produced stable and biologically plausible causal estimates while explicitly decoding pleiotropic instruments. In the BMI-T2D analysis, the decoded loci were corroborated by colocalisation (48% supported a shared causal variant, including *TCF7L2* at PP.H4 = 0.986) and by phenome-wide association patterns that separated an insulin-secretion channel (*TCF7L2*) from adiposity-driven pleiotropy (*FTO*). This illustrates how ECC-MR can enhance both interpretability and efficiency in settings where pleiotropy and LD are pervasive. Starting purely from statistical inconsistency among correlated instruments, the decoder recovered loci enriched for known antidiabetic and anti-obesity drug targets (*KCNJ11, PPARG* and *GIPR*), which was an independent, pharmacology-level validation of the decoding step. The other two applications illustrate ECC-MR’s behaviour at the lower end of the pleiotropic-burden gradient. For IOP-glaucoma, a pair with weak pleiotropy, ECC-MR retained all 1,360 correlated instruments and flagged no pleiotropic variant. Its estimate (θ□ = 1.249, 95% CI 1.034-1.454) was concordant with LD-pruned IVW, and the LD-block bootstrap kept the standard error honest rather than spuriously narrow; the regime in which retaining correlated instruments is pure gain, since redundancy is exploited without any decoding penalty. For LDL-CAD, a moderately pleiotropic pair, the decoder concentrated 13 pleiotropic variants at the *SH2B3* locus, and colocalisation confirmed shared causal variants at both *SH2B3* (PP.H4 = 0.975) and *TOMM40/APOE* (PP.H4 = 0.998), with no locus flagged as an LD artefact. The *SH2B3* finding is biologically coherent: its decoded variants associate with haematologic and inflammatory traits beyond lipids, pointing to a non-lipid channel through which LDL-associated loci may influence coronary risk, and echoing the locus’s known role as a pleiotropy hotspot in cardiovascular and blood traits. Notably, the strength of colocalisation evidence tracked the pleiotropic burden across the three pairs, i.e. no decoded variants in IOP–glaucoma, a compact confirmed set in LDL-CAD and a mixed picture of genuine pleiotropy and LD artefacts in BMI-T2D, suggesting that the decoder’s output composes a graded, verifiable map of pleiotropic architecture rather than a binary verdict.

Several limitations merit consideration. ECC-MR relies on sparsity assumptions regarding pleiotropy and requires accurate LD estimation from appropriate reference panels. Extensions to accommodate widespread directional pleiotropy or multiple causal pathways warrant further investigation. Future work may also explore adaptations of ECC-MR to multivariable and nonlinear MR settings.

In summary, ECC-MR provides a principled, interpretable and robust framework for causal inference in genetic epidemiology. By bridging Mendelian randomization and error-correcting codes, ECC-MR opens new avenues for leveraging redundancy and structure in genetic data to improve causal inference. By decoding pleiotropic variants rather than discarding them, ECC-MR turns pleiotropy into a starting point for mechanistic discovery, linking statistical signals directly to biology.

## Acknowledgementss

The authors thank the IEU OpenGWAS Project, the International Glaucoma Genetics Consortium, UK Biobank and FinnGen projects for making summary data publicly available.

## Author Contributions

J.J., D.H. and Q.Z. downloaded and analysed the data; J.J., D.H., Q.Z., X.L., S.Q., Q.L., M.T., S.S., Y.L., X.L. and Q.X. wrote the manuscript. Z.L. conceived and designed this study. G.C., L.D. and Z.L. supervised this work and reviewed the manuscript. All authors interpreted results and approved the manuscript.

## Ethical approval

All data used were publicly available, and no ethical approval was needed for this work.

## Data Availability Statement

The publicly available GWAS data are publicly available through the official portal (https://opengwas.io/datasets/). The code used in this work and R package ‘eccmr’ is available via GitHub at https://github.com/DrWoodWood/ECCMR.

## References

1. Smith, G.D. and S. Ebrahim, ‘Mendelian randomization’: can genetic epidemiology contribute to understanding environmental determinants of disease? Int J Epidemiol, 2003. 32(1): p. 1–22.

2. Lawlor, D.A., et al., Mendelian randomization: using genes as instruments for making causal inferences in epidemiology. Stat Med, 2008. 27(8): p. 1133–63.

3. Burgess, S., A. Butterworth, and S.G. Thompson, Mendelian randomization analysis with multiple genetic variants using summarized data. Genet Epidemiol, 2013. 37(7): p. 658–65.

4. Bowden, J., G. Davey Smith, and S. Burgess, Mendelian randomization with invalid instruments: effect estimation and bias detection through Egger regression. Int J Epidemiol, 2015. 44(2): p. 512–25.

5. Bowden, J., et al., Consistent Estimation in Mendelian Randomization with Some Invalid Instruments Using a Weighted Median Estimator. Genet Epidemiol, 2016. 40(4): p. 304–14.

6. Burgess, S., et al., Sensitivity Analyses for Robust Causal Inference from Mendelian Randomization Analyses with Multiple Genetic Variants. Epidemiology, 2017. 28(1): p. 30–42.

7. Hartwig, F.P., G. Davey Smith, and J. Bowden, Robust inference in summary data Mendelian randomization via the zero modal pleiotropy assumption. Int J Epidemiol, 2017. 46(6): p. 1985–1998.

8. Verbanck, M., et al., Detection of widespread horizontal pleiotropy in causal relationships inferred from Mendelian randomization between complex traits and diseases. Nat Genet, 2018. 50(5): p. 693–698.

9. Burgess, S., et al., Mendelian randomization with fine-mapped genetic data: Choosing from large numbers of correlated instrumental variables. Genet Epidemiol, 2017. 41(8): p. 714–725.

10. Zhu, Z., et al., Integration of summary data from GWAS and eQTL studies predicts complex trait gene targets. Nat Genet, 2016. 48(5): p. 481–7.

11. Donoho, D.L., Compressed sensing. IEEE Transactions on Information Theory, 2006. 52(4): p. 1289–1306.

12. Candès, E.J., J.K. Romberg, and T. Tao, Stable signal recovery from incomplete and inaccurate measurements. Communications on Pure and Applied Mathematics, 2006. 59(8): p. 1207–1223.

13. Berisa, T. and J.K. Pickrell, Approximately independent linkage disequilibrium blocks in human populations. Bioinformatics, 2016. 32(2): p. 283–5.

14. Tseng, P., Convergence of a Block Coordinate Descent Method for Nondifferentiable Minimization. Journal of Optimization Theory and Applications, 2001. 109(3): p. 475–494.

15. Candès, E.J., The restricted isometry property and its implications for compressed sensing. Comptes Rendus Mathematique, 2008. 346(9): p. 589–592.

16. Verbanck, M., et al., Detection of widespread horizontal pleiotropy in causal relationships inferred from Mendelian randomization between complex traits and diseases. Nature Genetics, 2018. 50(5): p. 693–698.

17. Rees, J.M.B., et al., Robust methods in Mendelian randomization via penalization of heterogeneous causal estimates. PLOS ONE, 2019. 14(9): p. e0222362.

18. Qi, G. and N. Chatterjee, Mendelian randomization analysis using mixture models for robust and efficient estimation of causal effects. Nature Communications, 2019. 10(1): p. 1941.

19. Xue, H., X. Shen, and W. Pan, Constrained maximum likelihood-based Mendelian randomization robust to both correlated and uncorrelated pleiotropic effects. Am J Hum Genet, 2021. 108(7): p. 1251–1269.

20. Zhu, Z., et al., Causal associations between risk factors and common diseases inferred from GWAS summary data. Nature Communications, 2018. 9(1): p. 224.

21. Foley, C.N., et al., MR-Clust: clustering of genetic variants in Mendelian randomization with similar causal estimates. Bioinformatics, 2021. 37(4): p. 531–541.

22. Willer, C.J., et al., Discovery and refinement of loci associated with lipid levels. Nat Genet, 2013. 45(11): p. 1274–1283.

23. Nikpay, M., et al., A comprehensive 1,000 Genomes-based genome-wide association meta-analysis of coronary artery disease. Nat Genet, 2015. 47(10): p. 1121–1130.

24. Bycroft, C., et al., The UK Biobank resource with deep phenotyping and genomic data. Nature, 2018. 562(7726): p. 203–209.

25. Kurki, M.I., et al., FinnGen provides genetic insights from a well-phenotyped isolated population. Nature, 2023. 613(7944): p. 508–518.

26. Elsworth, B., et al., The MRC IEU OpenGWAS data infrastructure. bioRxiv, 2020: p. 2020.08.10.244293.

