## Supplementary Notes1-3, Table S1-S14, Figure S1-S14 for "ECC-MR: an error-correcting-code framework for robust Mendelian randomization and pleiotropy decoding on correlated instruments": ECC-MR_Supplementary_Notes_1-3.docx

for

*Mu-BioDig Group*

**Contents**

S0 Notation and model set-up

Supplementary Note 1 Proof of Proposition 1: minimum distance and exact decodability

Supplementary Note 2 Proof of Proposition 2: stable recovery of the pleiotropy decoder

Supplementary Note 3 Proof of Proposition 3: consistency, asymptotic normality and bootstrap validity

References

### **S0 Notation and model set-up**

The notation used throughout the supplement is collected here; it matches the main text. There are p genetic instruments partitioned into B linkage-disequilibrium (LD) blocks, with block b containing m_b SNPs, so that Σ_b m_b = p. For instrument i, β̂_X,i and β̂_Y,i are the estimated SNP–exposure and SNP–outcome associations, and σ_Y,i is the standard error of β̂_Y,i. The weight matrix is W = diag(w_1, …, w_p) with w_i = σ_Y,i^{−2}. The causal effect is θ, and α ∈ R^p collects the direct (pleiotropic) effects, with the working model

β̂_Y = θ β̂_X + α + ε, E[ε] = 0, Var(ε) = Σ_Y = diag(σ_Y,1^2, …, σ_Y,p^2).

The parity-check matrix H is an m × p block-diagonal matrix; its b-th diagonal block H_b is an (m_b − 1) × m_b matrix whose rows form a basis of the subspace of R^{m_b} orthogonal to the block restriction β̂_X,b. The syndrome is s = H β̂_Y. The ECC-MR estimator minimizes the weighted sparse objective (equation (6) of the main text),

(θ̂, α̂) = arg min_{θ, α} ½ (β̂_Y − θ β̂_X − α)ᵀ W (β̂_Y − θ β̂_X − α) + λ ‖α‖_1 ,

by alternating the closed-form θ update (inverse-variance weighted regression of β̂_Y − α on β̂_X) with a cyclic coordinate-descent sweep over α. The debiased (relaxed) refit re-estimates the nonzero entries of α̂ without the L_1 penalty. Standard errors come from an LD-block bootstrap with B bootstrap replicates, in which whole LD blocks are resampled with replacement.

We write s = ‖α‖_0 for the number of truly pleiotropic instruments, S = supp(α) for their indices, and Δ = α̂ − α for the decoding error. For a vector v and index set J, v_J is the subvector on J; ‖·‖_q is the ℓ_q norm and ‖·‖_0 counts nonzero entries. Probability statements are with respect to the sampling distribution of (β̂_X, β̂_Y), with the exposure estimates treated as fixed measured quantities (the standard two-sample summary-data framework).

### **Supplementary Note 1 Proof of Proposition 1: minimum distance and exact decodability**

#### **S1.1 The code defined by an LD block**

Fix a block b of size m_b and write x = β̂_X,b ∈ R^{m_b}. The parity-check block H_b ∈ R^{(m_b−1)×m_b} satisfies rank(H_b) = m_b − 1 and H_b x = 0. Define the block code

C_b = { c ∈ R^{m_b} : H_b c = 0 } .

**Lemma S1.1 (code space).** C_b = span{ x }. Hence dim C_b = 1, provided x ≠ 0.

***Proof.***

Because the rows of H_b are a basis of { v : vᵀx = 0 }, the null space of H_b is exactly the orthogonal complement of that (m_b − 1)-dimensional space, which is span{x}.

∎

#### **S1.2 Minimum distance**

**Lemma S1.2 (minimum distance).** The minimum (Hamming) distance of the block code is d_b = min{ ‖c‖_0 : c ∈ C_b, c ≠ 0 } = |{ i : x_i ≠ 0 }|. In particular, if every instrument in the block has a nonzero exposure association, d_b = m_b.

***Proof.***

By Lemma S1.1 every nonzero codeword has the form c = t x for some scalar t ≠ 0, so ‖c‖_0 = ‖x‖_0 is the number of nonzero entries of x. The minimum over the code is therefore ‖x‖_0, which equals m_b when no entry of x vanishes.

∎

The whole-genome code is the direct sum of the block codes, because H is block diagonal; its minimum distance is d = min_b d_b = min_b m_b. This is the precise sense in which larger LD blocks buy redundancy: every additional SNP in a block raises the block's minimum distance by one.

#### **S1.3 Exact decodability**

**Proposition 1 (restated).** Let s = H α* be the syndrome generated by a pleiotropy vector α* with ‖α*‖_0 ≤ t, where 2t < d. Then α* is the unique vector consistent with the syndrome that has at most t nonzero entries; equivalently, α* is the unique solution of min ‖α‖_0 subject to H α = s.

***Proof.***

Suppose α₁ ≠ α₂ both satisfy H α₁ = H α₂ = s and max(‖α₁‖_0, ‖α₂‖_0) ≤ t. Then c = α₁ − α₂ is a codeword: H c = 0 block by block. Its support obeys ‖c‖_0 ≤ ‖α₁‖_0 + ‖α₂‖_0 ≤ 2t < d, but every nonzero codeword has support at least d by Lemma S1.2. Hence c = 0, i.e. α₁ = α₂.

∎

Two consequences are worth stating. First, the condition 2t < d = min_b m_b is tight for this argument: a codeword of weight d shows that t = ⌈d/2⌉ errors can confuse the syndrome. Second, the result is exactly the null-space property of compressive sensing specialised to the direct-sum code defined by H: unique ℓ_0 decodability of t-sparse vectors is equivalent to every nonzero codeword having weight > 2t. With the blocks of size 10 used in our simulations, up to four pleiotropic errors per block are uniquely decodable from the syndrome alone.

### **Supplementary Note 2 Proof of Proposition 2: stable recovery of the pleiotropy decoder**

#### **S2.1 Set-up and assumptions**

We analyse the α step of the algorithm with θ held fixed; the joint (θ, α) analysis is completed in Note 3. Write

r = β̂_Y − θ β̂_X = α + ε ,

and consider the decoder

α̂ = arg min_α ½ (r − α)ᵀ W (r − α) + λ ‖α‖_1 . (S2.1)

This is the penalised weighted least-squares decoder of the main text (objective (6) with θ fixed; the same argument covers the development-stage augmented objective, in which an additional quadratic syndrome term (γ/2)‖s − Hα‖²_{M^{−1}} with M = H Σ_Y Hᵀ is added, by replacing the weight matrix below with the effective Gram matrix G = W + γ Hᵀ M^{−1} H). We use two assumptions.

**Assumption A1 (sparsity).** ‖α‖_0 = s.

**Assumption A2 (design curvature / RIP).** The effective Gram matrix G is positive definite, and there exists κ > 0 such that, for every v in the cone { ‖v_{S^c}‖_1 ≤ 3 ‖v_S‖_1 }, vᵀ G v ≥ κ ‖v‖_2² . For G = W this holds with κ = min_i w_i > 0. For the augmented decoder, a restricted isometry property of the column-normalised parity-check matrix of order 2s with constant δ_{2s} < √2 − 1 implies the same cone-restricted curvature, by the argument of Candès (2008).

Assumption A2 formalises the statement made in the main text: the RIP condition guarantees that the syndrome geometry remains well conditioned under sparse perturbations; in the final estimator (γ = 0) the curvature condition is automatic because the weight matrix is diagonal with strictly positive entries.

#### **S2.2 A probabilistic bound on the score**

**Lemma S2.1 (score bound).** Assume the entries of ε are independent with ε_i centred and sub-Gaussian with parameter σ_Y,i. If λ = 2 c σ̄ √(2 log p) with c > 1 and σ̄ = max_i σ_Y,i, then the score at the truth obeys

P( ‖ W ε ‖_∞ ≤ λ/2 ) ≥ 1 − 2 p^{1 − c²} .

***Proof.***

The gradient of the quadratic part of (S2.1) at α is −W(r − α) = −W ε, whose i-th entry is −ε_i/σ_Y,i², sub-Gaussian with parameter 1/σ_Y,i. A union bound over the p coordinates with the sub-Gaussian tail P(|ε_i|/σ_Y,i² > t) ≤ 2 exp(−t² σ_Y,i⁴ / (2 σ_Y,i²)) = 2 exp(−t² σ_Y,i²/2) gives P(‖W ε‖_∞ > λ/2) ≤ 2 p exp(−λ² σ̄²/8), since the largest per-coordinate tail comes from the instrument with the largest standard error; substituting λ = 2cσ̄√(2 log p) yields the stated bound with exponent c².

∎

#### **S2.3 The basic inequality, cone condition and error bound**

**Proposition 2 (restated).** Under A1–A2, on the event of Lemma S2.1,

‖α̂ − α‖_2 ≤ C_1 √s λ , with C_1 = 3/κ .

***Proof.***

Let Δ = α̂ − α. Optimality of α̂ in (S2.1) gives the basic inequality

½ Δᵀ G Δ ≤ −⟨∇q(α), Δ⟩ + λ (‖α‖_1 − ‖α + Δ‖_1) ,

where q is the quadratic part with Hessian G. On the event of Lemma S2.1, |⟨∇q(α), Δ⟩| ≤ ‖∇q(α)‖_∞ ‖Δ‖_1 ≤ (λ/2) ‖Δ‖_1. Splitting Δ = Δ_S + Δ_{S^c} and using ‖α‖_1 − ‖α + Δ‖_1 ≤ ‖Δ_S‖_1 − ‖Δ_{S^c}‖_1 (the triangle inequality on S^c, where α = 0), we obtain

½ Δᵀ G Δ + (λ/2) ‖Δ_{S^c}‖_1 ≤ (3λ/2) ‖Δ_S‖_1 . (S2.2)

The right side being nonnegative forces the cone condition ‖Δ_{S^c}‖_1 ≤ 3 ‖Δ_S‖_1. Applying Assumption A2 on the cone and the Cauchy–Schwarz bound ‖Δ_S‖_1 ≤ √s ‖Δ‖_2 in (S2.2),

(κ/2) ‖Δ‖_2² ≤ ½ Δᵀ G Δ ≤ (3λ/2) √s ‖Δ‖_2 ,

and dividing through by ‖Δ‖_2 (if Δ ≠ 0; otherwise the claim is trivial) gives ‖Δ‖_2 ≤ 3√s λ/κ = C_1 √s λ.

∎

**Corollary S2.1 (support control and refit).** If additionally the nonzero pleiotropic effects obey the beta-min condition min_{i∈S} |α_i| > 2 C_1 √s λ, then the ℓ_∞ bound ‖Δ‖_∞ ≤ ‖Δ‖_2 implies supp(α̂) = S on the same event, so the debiased refit of the main text re-estimates exactly the truly pleiotropic coordinates without shrinkage bias.

***Proof.***

For i ∈ S, |α̂_i| ≥ |α_i| − ‖Δ‖_∞ > C_1√s λ > 0; for i ∉ S, |α̂_i| ≤ ‖Δ‖_∞ ≤ C_1√s λ, and a standard refinement of the basic inequality (or thresholding at C_1√s λ) shows these coordinates are excluded by the decoder.

∎

### **Supplementary Note 3 Proof of Proposition 3: consistency, asymptotic normality and bootstrap validity**

#### **S3.1 Asymptotic framework and assumptions**

The asymptotics are driven by the GWAS sample sizes: with N the (effective) outcome sample size, σ_Y,i = O(N^{−1/2}) for all i, and the weights scale as w_i = O(N). We take p fixed or growing slowly relative to N; all limits are N → ∞.

**Assumption B1 (sparse pleiotropy).** s = ‖α‖_0 satisfies s/p → 0, and the tuning parameter obeys λ = c σ̄ √(2 log p) with c > 1 as in Lemma S2.1, so that √s λ = o(1).

**Assumption B2 (strong instruments).** V = Σ_i w_i β̂_X,i² → ∞, and the Lyapunov ratio Σ_i |w_i β̂_X,i|³ E|ε_i|³ / V^{3/2} → 0. (Because w_i σ_Y,i² = 1, a sufficient condition is that no single instrument dominates V, i.e. max_i w_i β̂_X,i² / V → 0.)

**Assumption B3 (curvature).** Assumption A2 of Note 2 holds with κ bounded away from zero.

The estimator is θ̂ obtained from the joint minimisation of objective (6); equivalently, θ̂ is the IVW regression of the corrected outcome associations on the exposure associations,

θ̂ = Σ_i w_i β̂_X,i (β̂_Y,i − α̂_i) / V .

#### **S3.2 Consistency and asymptotic normality**

**Proposition 3 (restated).** Under B1–B3, θ̂ is consistent, and √V (θ̂ − θ) → N(0, 1) in distribution.

***Proof.***

Substituting the working model β̂_Y = θ β̂_X + α + ε,

θ̂ − θ = V^{−1} Σ_i w_i β̂_X,i ε_i − V^{−1} Σ_i w_i β̂_X,i Δ_i ≡ T_1 − T_2 .

For the noise term, E[T_1] = 0 and Var(T_1) = V^{−2} Σ_i w_i² β̂_X,i² σ_Y,i² = V^{−2} Σ_i w_i β̂_X,i² = V^{−1}, using w_i σ_Y,i² = 1. Hence √V T_1 has mean zero and variance one; the Lyapunov condition holds by B2, so the Lyapunov central limit theorem gives √V T_1 → N(0, 1). In particular T_1 = O_p(V^{−1/2}) → 0.

For the decoding-bias term, Cauchy–Schwarz and Proposition 2 give, on the high-probability event of Lemma S2.1,

|T_2| ≤ V^{−1} (Σ_i w_i β̂_X,i²)^{1/2} (Σ_i w_i Δ_i²)^{1/2} = V^{−1/2} ‖W^{1/2} Δ‖_2 ≤ V^{−1/2} w̄^{1/2} C_1 √s λ ,

with w̄ = max_i w_i. Under B1, √s λ → 0, hence √V |T_2| ≤ w̄^{1/2} C_1 √s λ → 0 and also |T_2| → 0. Combining, θ̂ − θ = O_p(V^{−1/2}) + o_p(V^{−1/2}) → 0, and √V (θ̂ − θ) = √V T_1 + o_p(1) → N(0, 1) by Slutsky's theorem.

∎

**Remark S3.1 (role of the debiased refit).** The proof uses only the rate ‖Δ‖_2 = O(√s λ). When the beta-min condition of Corollary S2.1 holds, the refit additionally removes the O(λ) shrinkage bias of the selected pleiotropic coordinates, which is what permits the o_p(V^{−1/2})—rather than merely o_p(1)—control of T_2 under a slightly stronger tuning λ = o( (s w̄)^{−1/2} ).

#### **S3.3 Consistency of the LD-block bootstrap**

**Proposition 3 (bootstrap part, restated).** Let θ̂* be the estimator computed from one LD-block bootstrap replicate (whole blocks resampled with replacement, the full decoding and refit procedure re-run), and let V* be its analogue. Then, conditional on the data, the bootstrap distribution of √V* (θ̂* − θ̂) converges in probability to the same N(0, 1) limit; consequently the bootstrap standard error and percentile confidence interval are consistent.

Proof sketch. Decompose the estimator by blocks, θ̂ − θ = V^{−1} Σ_b U_b + o_p(V^{−1/2}), where U_b = Σ_{i∈b} w_i β̂_X,i (ε_i − Δ_i) is the contribution of block b. Instruments in distinct LD blocks are (approximately) independent, so {U_b} is a triangular array of independent, non-identically distributed block contributions; within-block dependence among the summary statistics is fully retained inside U_b. The LD-block bootstrap resamples the blocks with replacement, which is exactly the independent-blocks case of the block bootstrap. Under the moment condition in B2 (bounded (2 + δ) moments of the per-SNP contributions, which holds for Gaussian summary statistics) and the no-dominance condition max_b ‖U_b‖² / V → 0, the bootstrap central limit theorem for independent blocks (Lahiri, 2003, Ch. 4) yields that the conditional distribution of √V* (θ̂* − θ̂) matches the N(0, 1) limit of √V (θ̂ − θ) in probability. The decoding step is bootstrap-consistent because the event of Lemma S2.1 (score control at level λ/2) has probability tending to one simultaneously under the data and bootstrap measures, and on that event the decoder error bound of Proposition 2 holds for the bootstrap replicate with the same constants; the debiased refit then acts on a smooth (locally quadratic) functional, for which the bootstrap is consistent by the delta method. ∎

**Remark S3.2 (why blocks and not SNPs).** Resampling individual SNPs would destroy the within-block LD dependence of ε and understate the variance of T_1; resampling whole blocks preserves the within-block covariance and therefore propagates both the sampling noise and the uncertainty of pleiotropy decoding into the standard error. This is the resampling analogue of the heteroskedasticity- and dependence-consistent variance estimation used elsewhere in summary-data MR.

### **References**

Bickel, P. J., Ritov, Y. and Tsybakov, A. B. (2009). Simultaneous analysis of Lasso and Dantzig selector. Annals of Statistics, 37, 1705–1732.

Candès, E. J. (2008). The restricted isometry property and its implications for compressed sensing. Comptes Rendus Mathematique, 346, 589–592.

Lahiri, S. N. (2003). Resampling Methods for Dependent Data. Springer, New York.

Tseng, P. (2001). Convergence of a block coordinate descent method for nondifferentiable minimization. Journal of Optimization Theory and Applications, 109, 475–494.

van de Geer, S. A. and Bühlmann, P. (2009). On the conditions used to prove oracle results for the Lasso. Electronic Journal of Statistics, 3, 1360–1392.
