## Supplementary figures and images for "ECC-MR: an error-correcting-code framework for robust Mendelian randomization and pleiotropy decoding on correlated instruments"

### Figure S1-S10.pdf

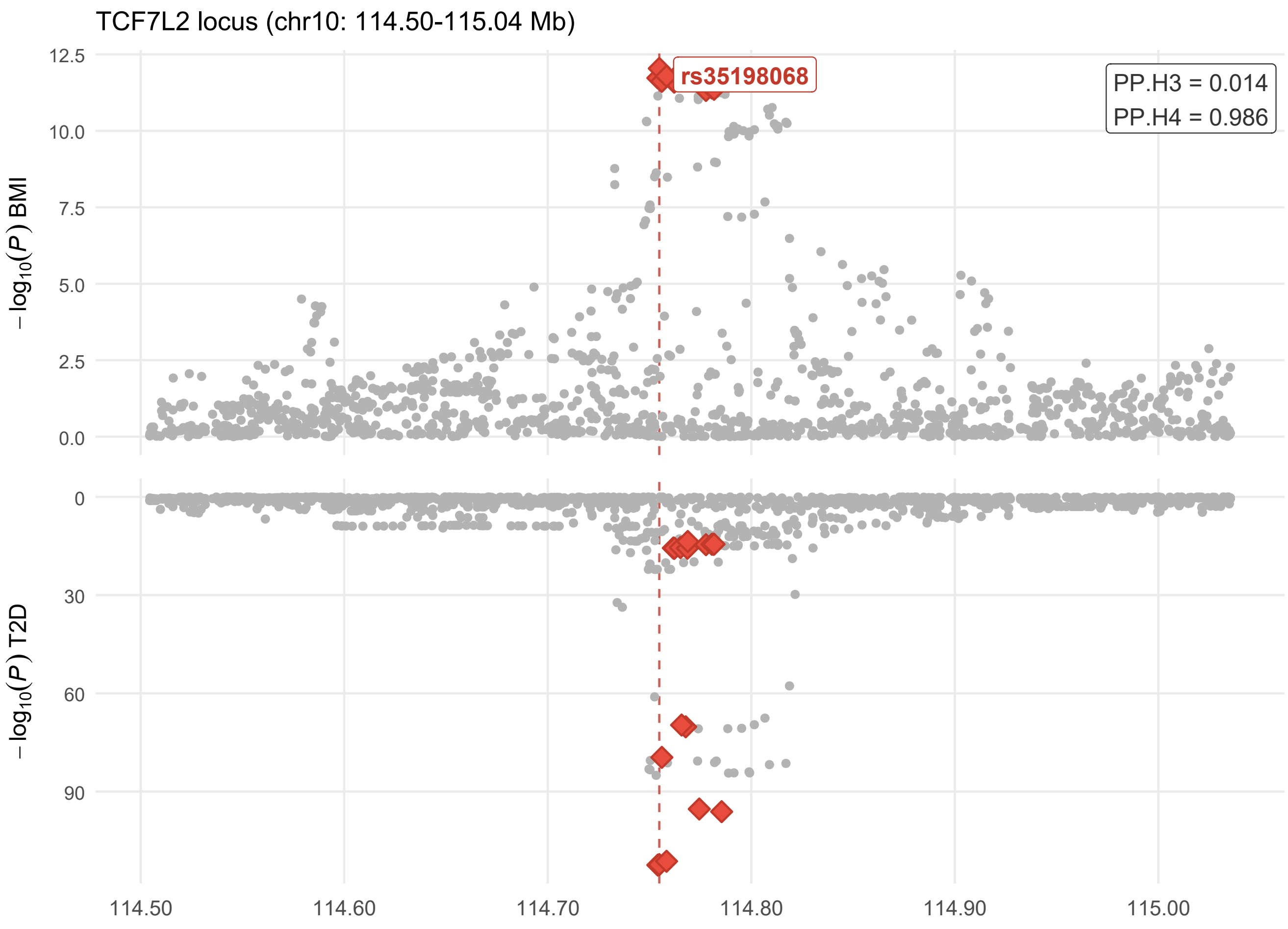

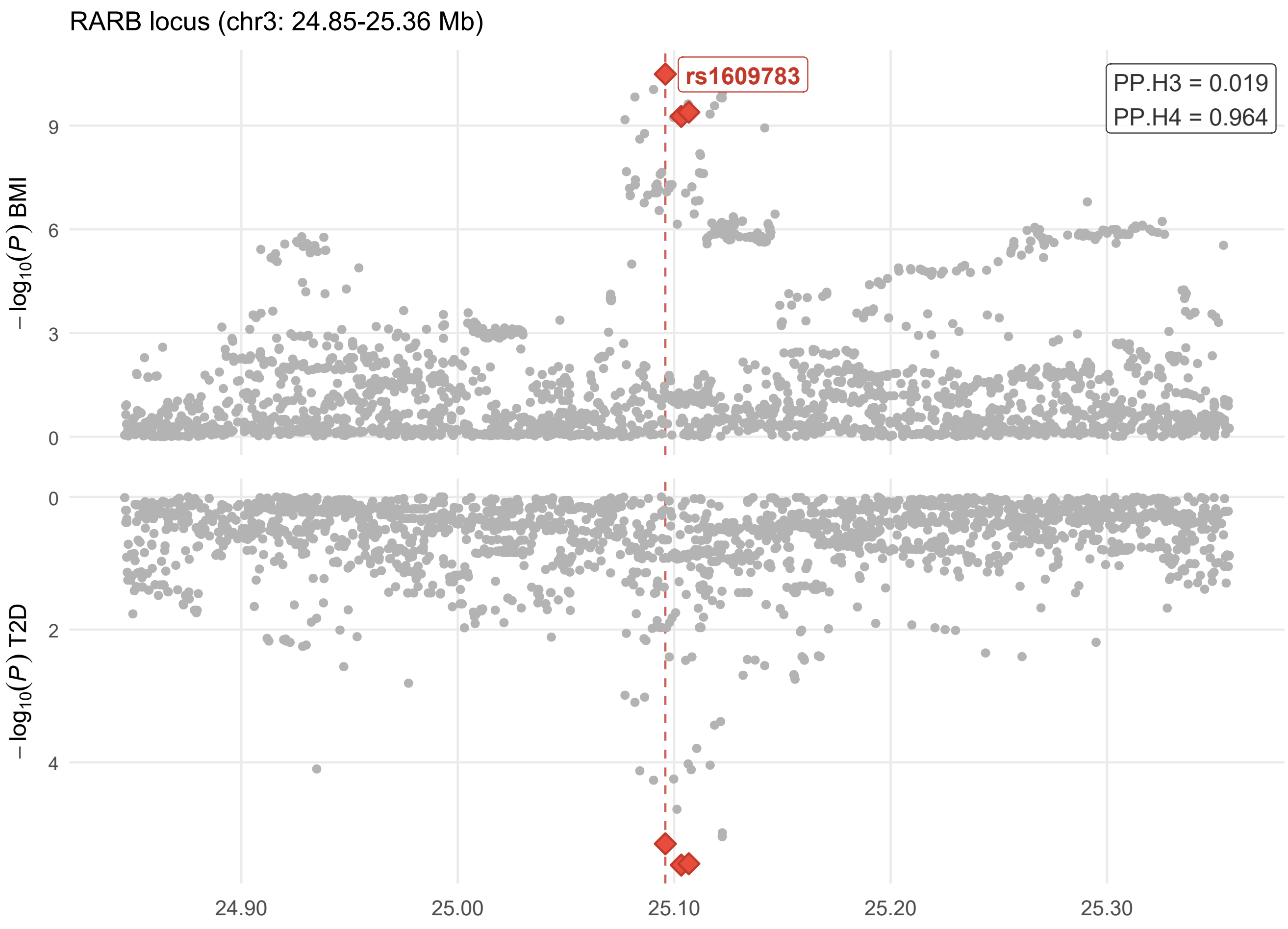

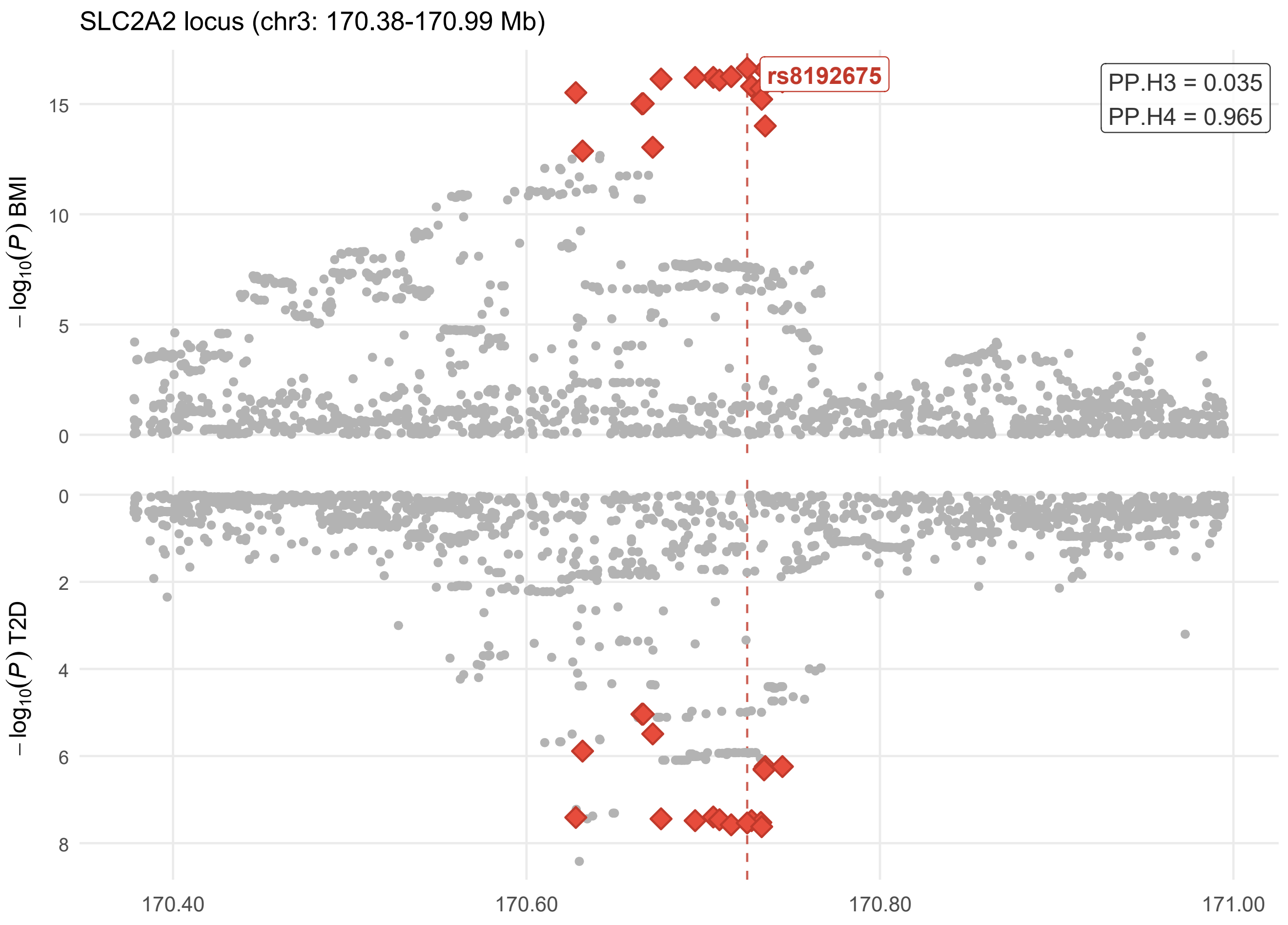

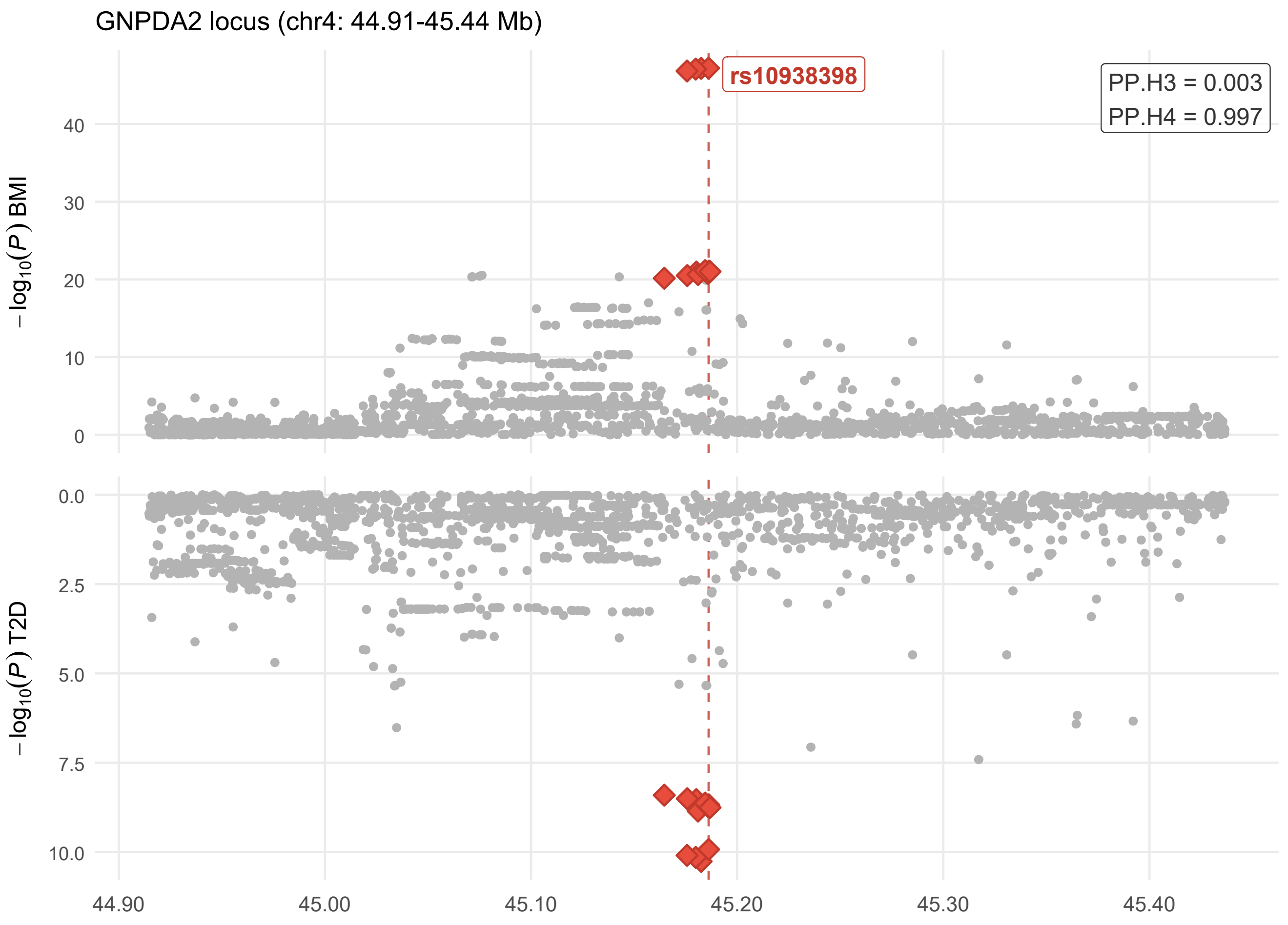

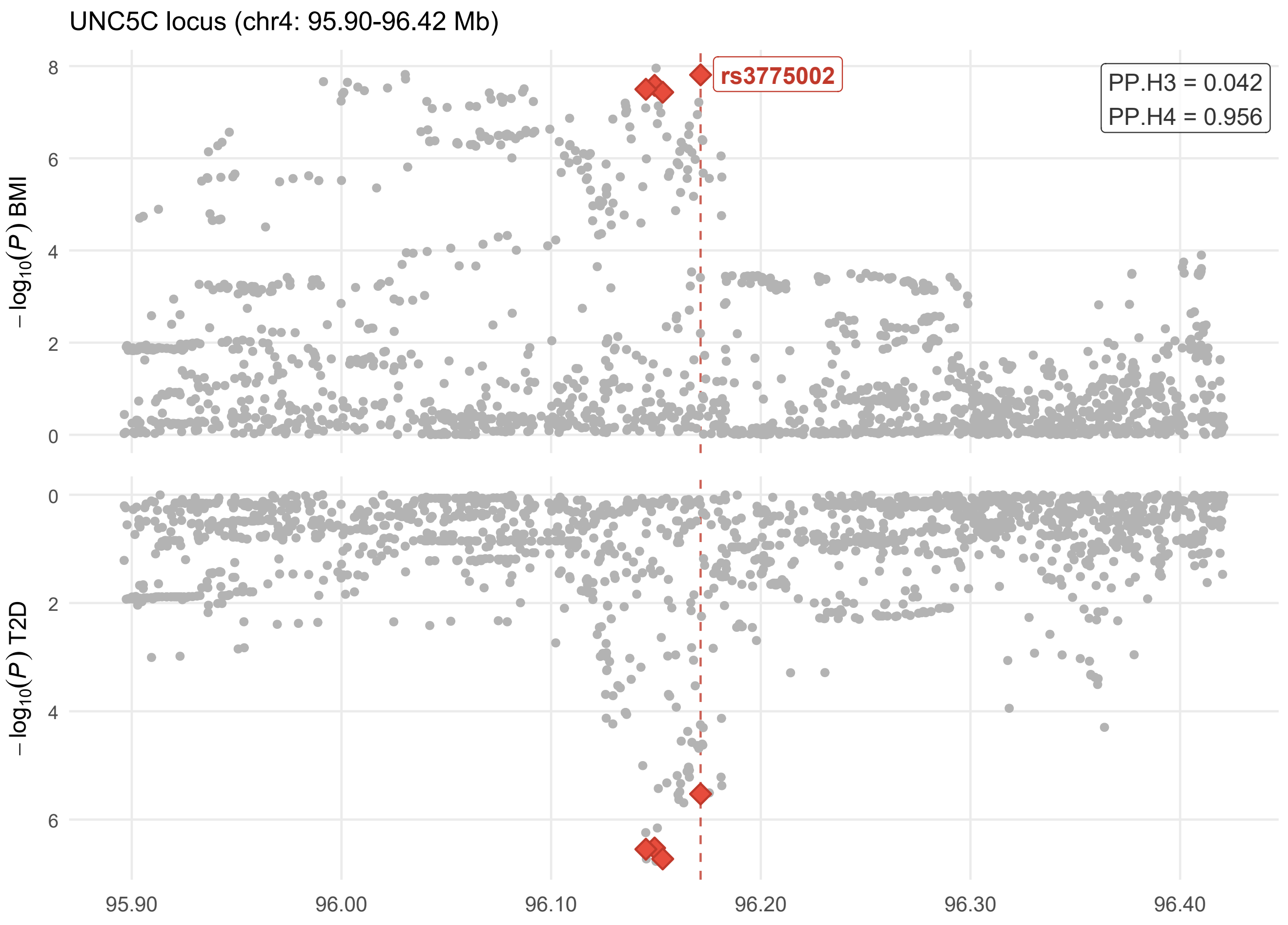

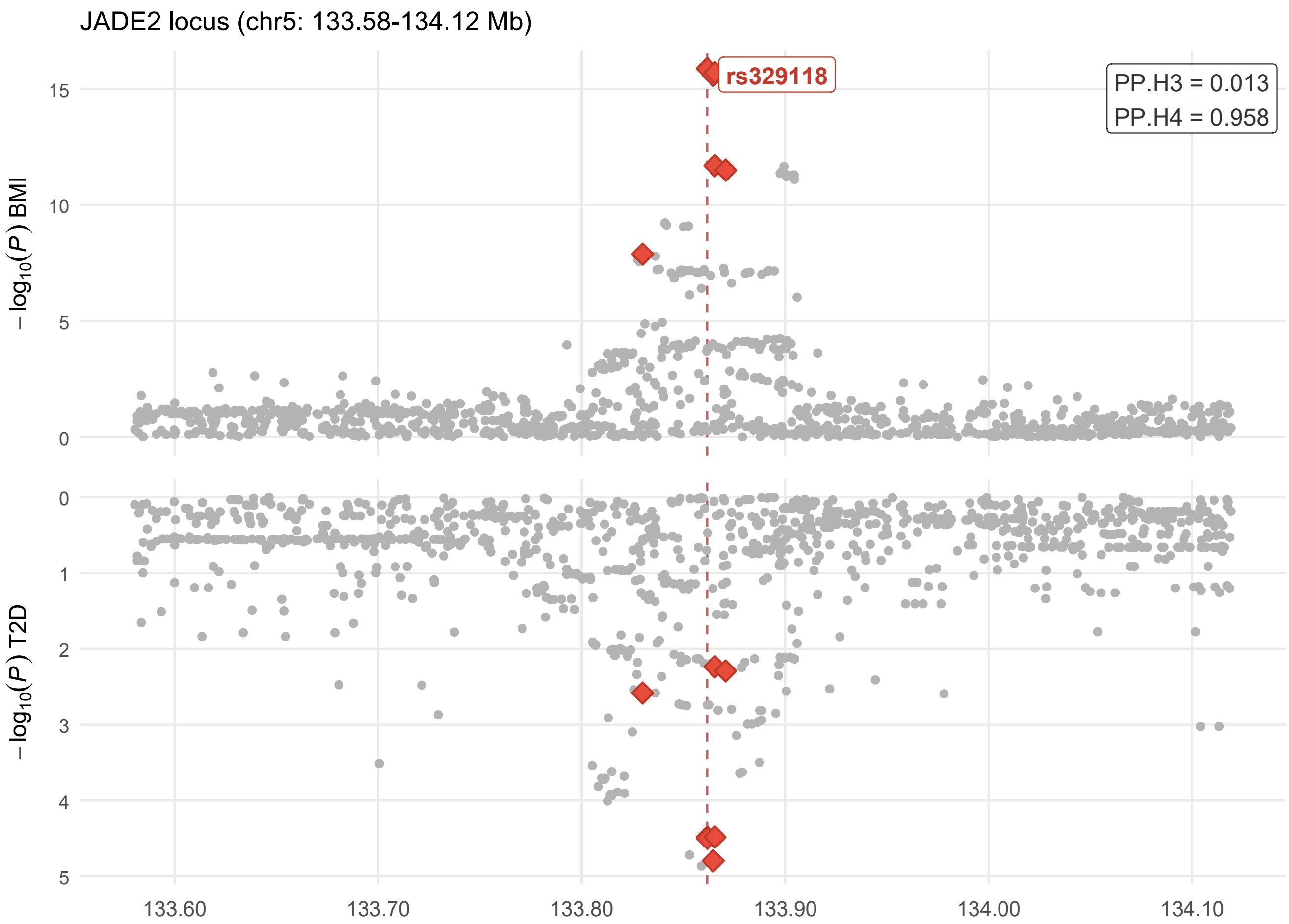

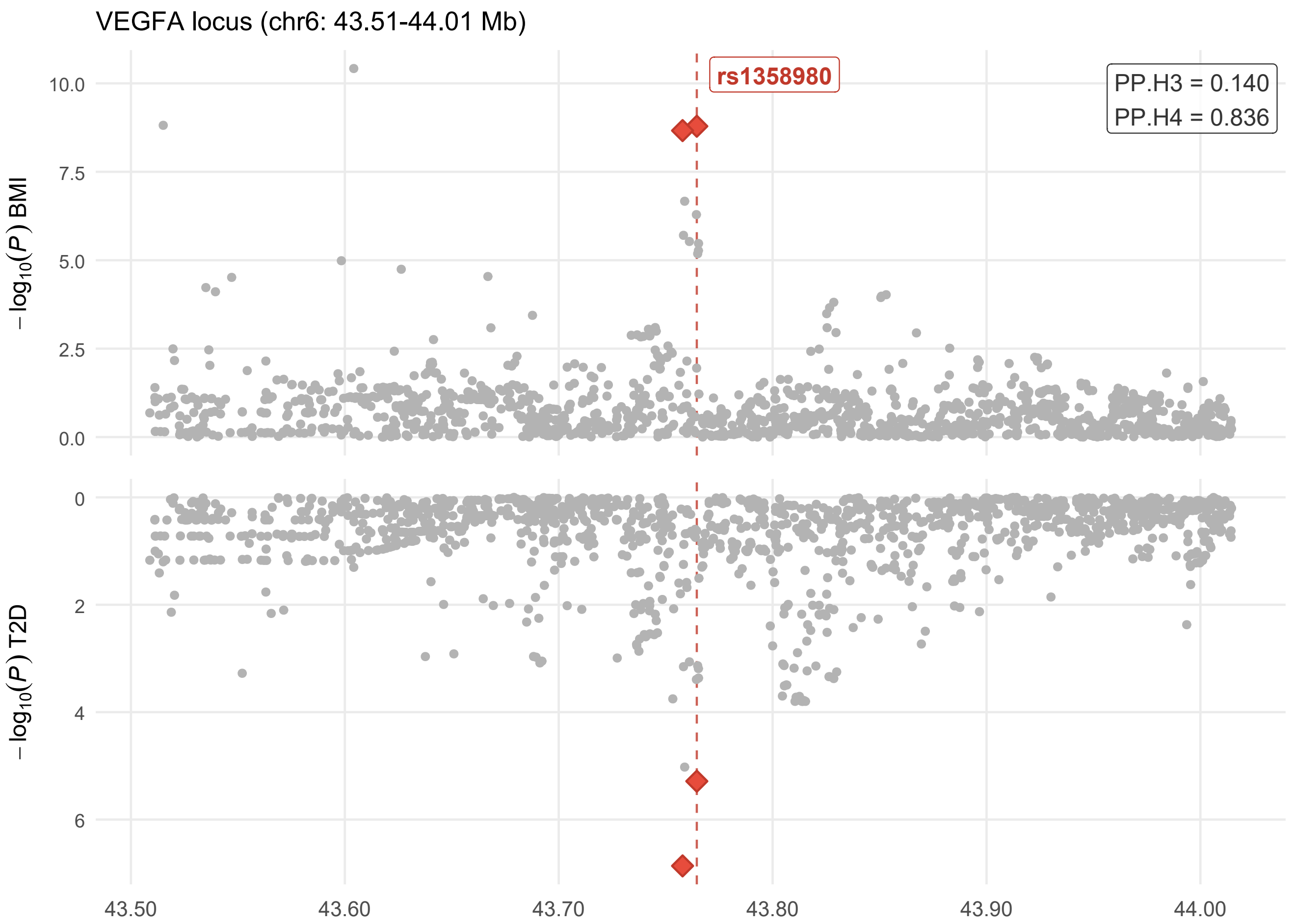

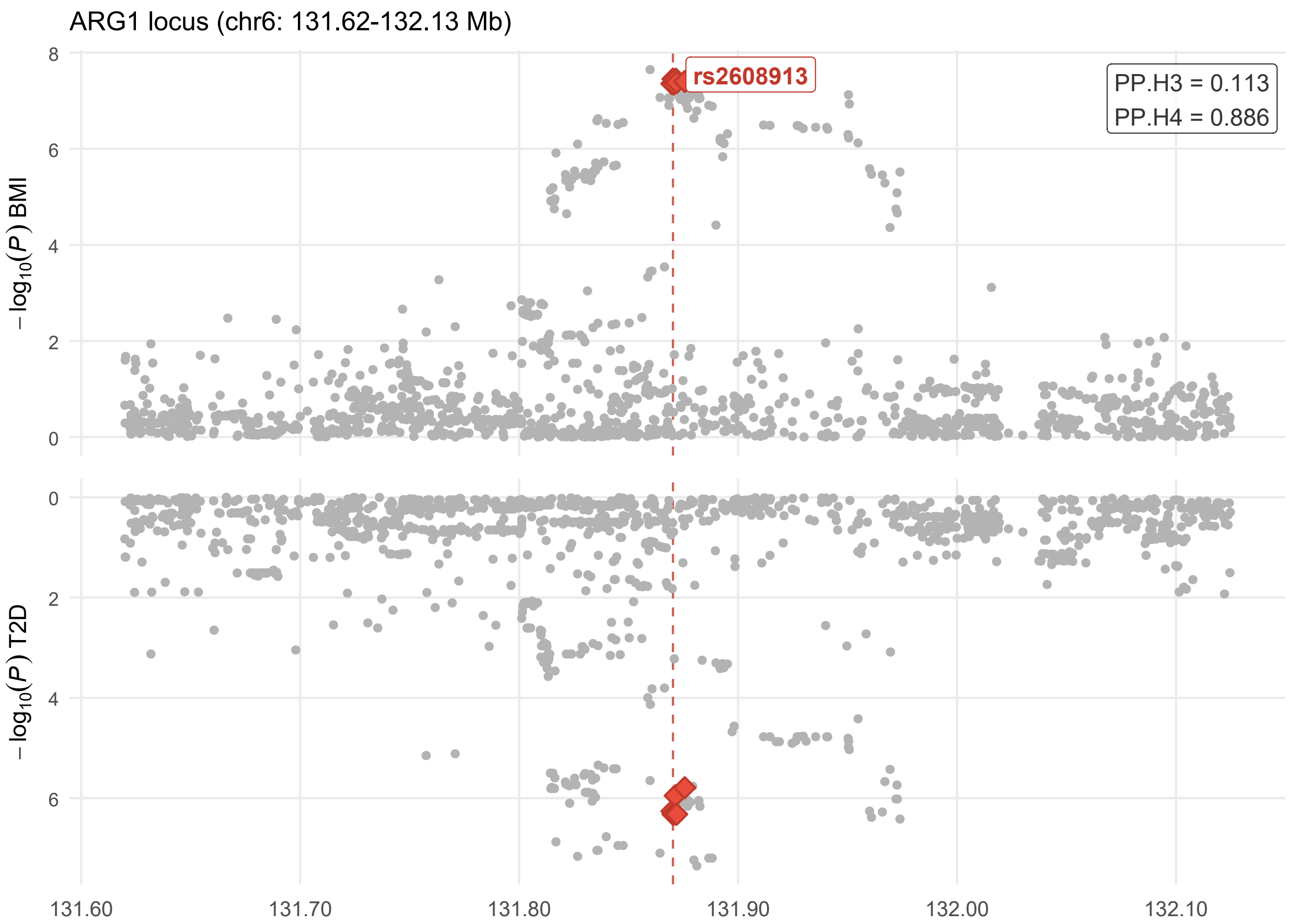

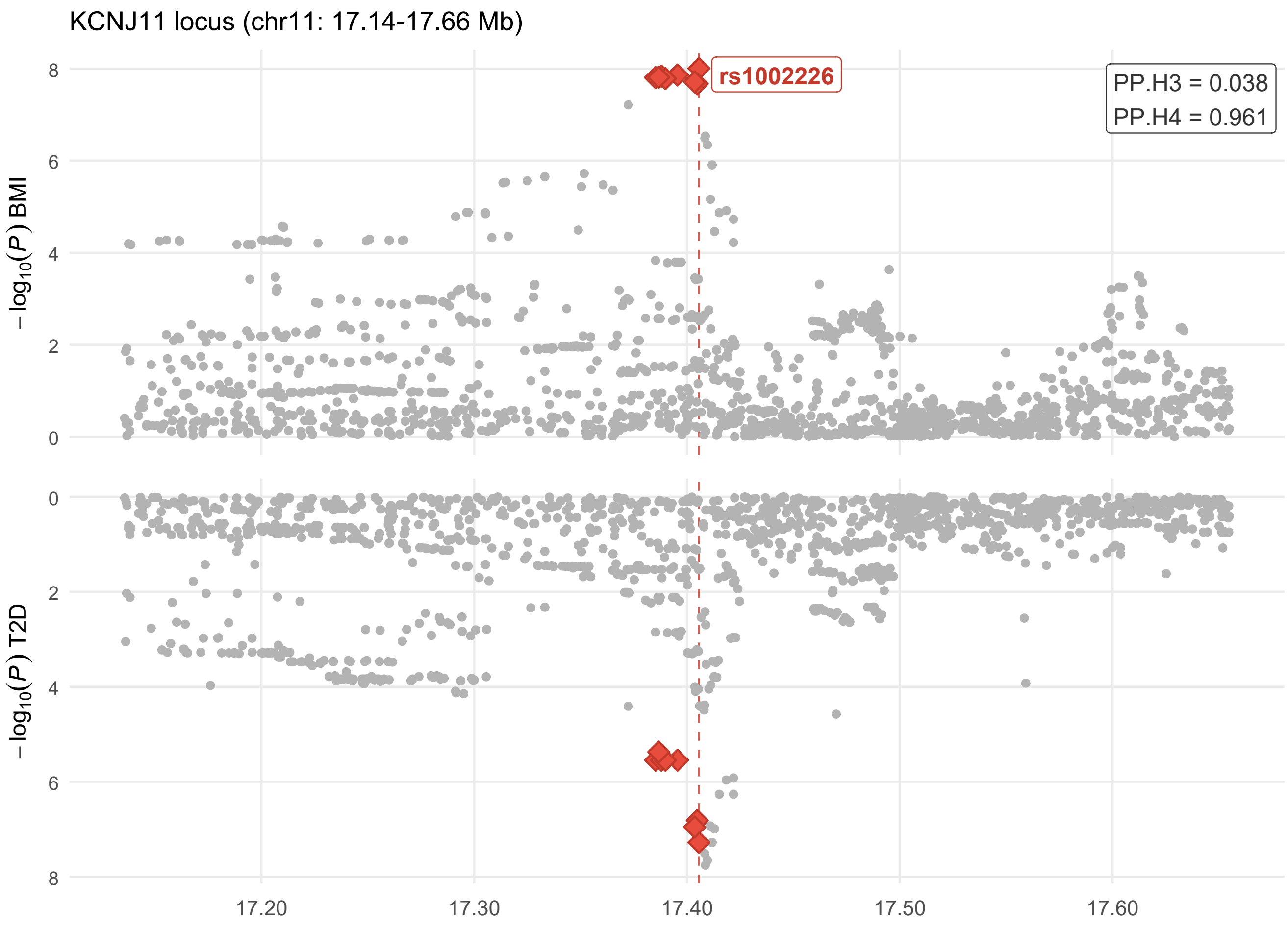

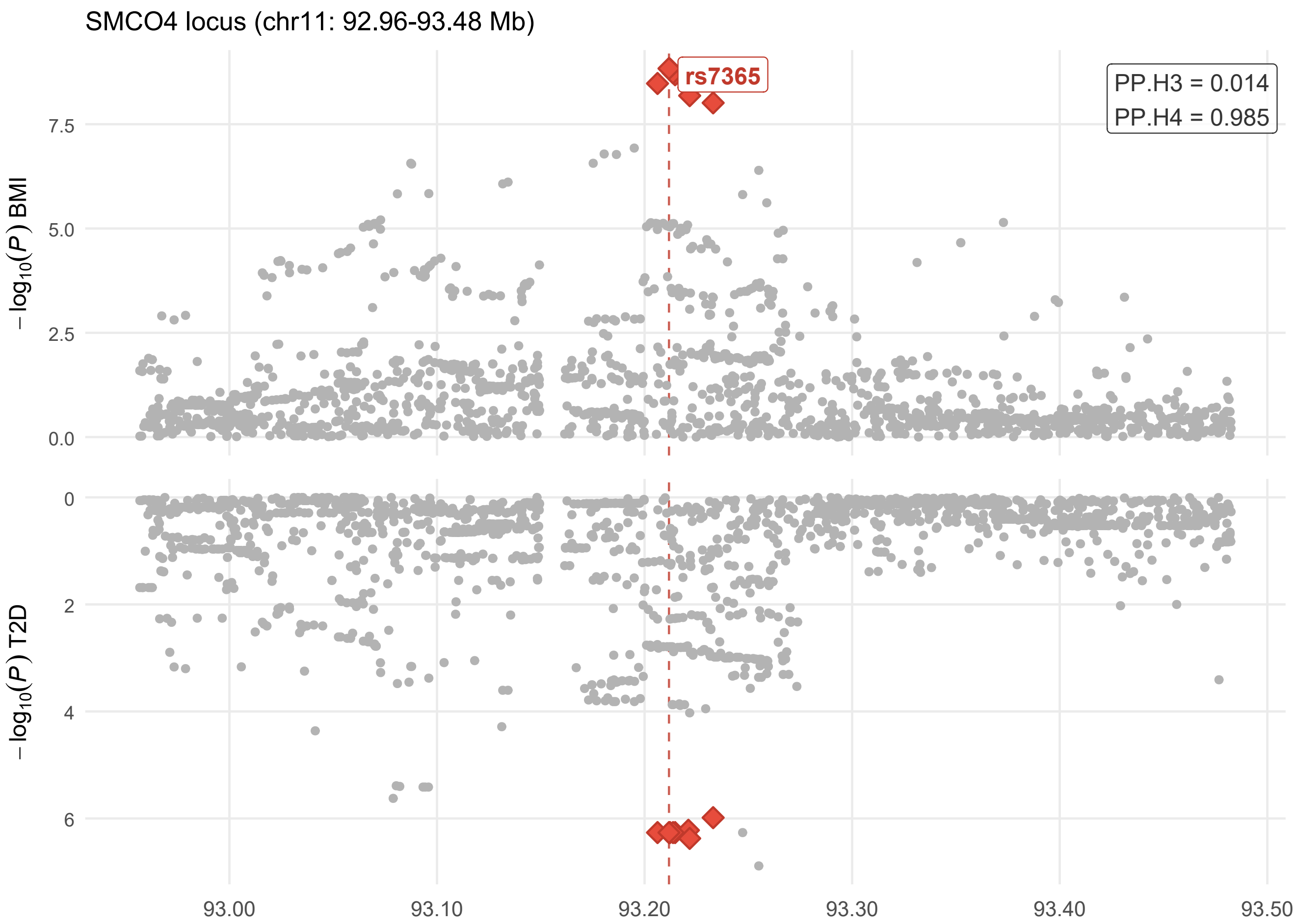

### Figure S11-S14.pdf

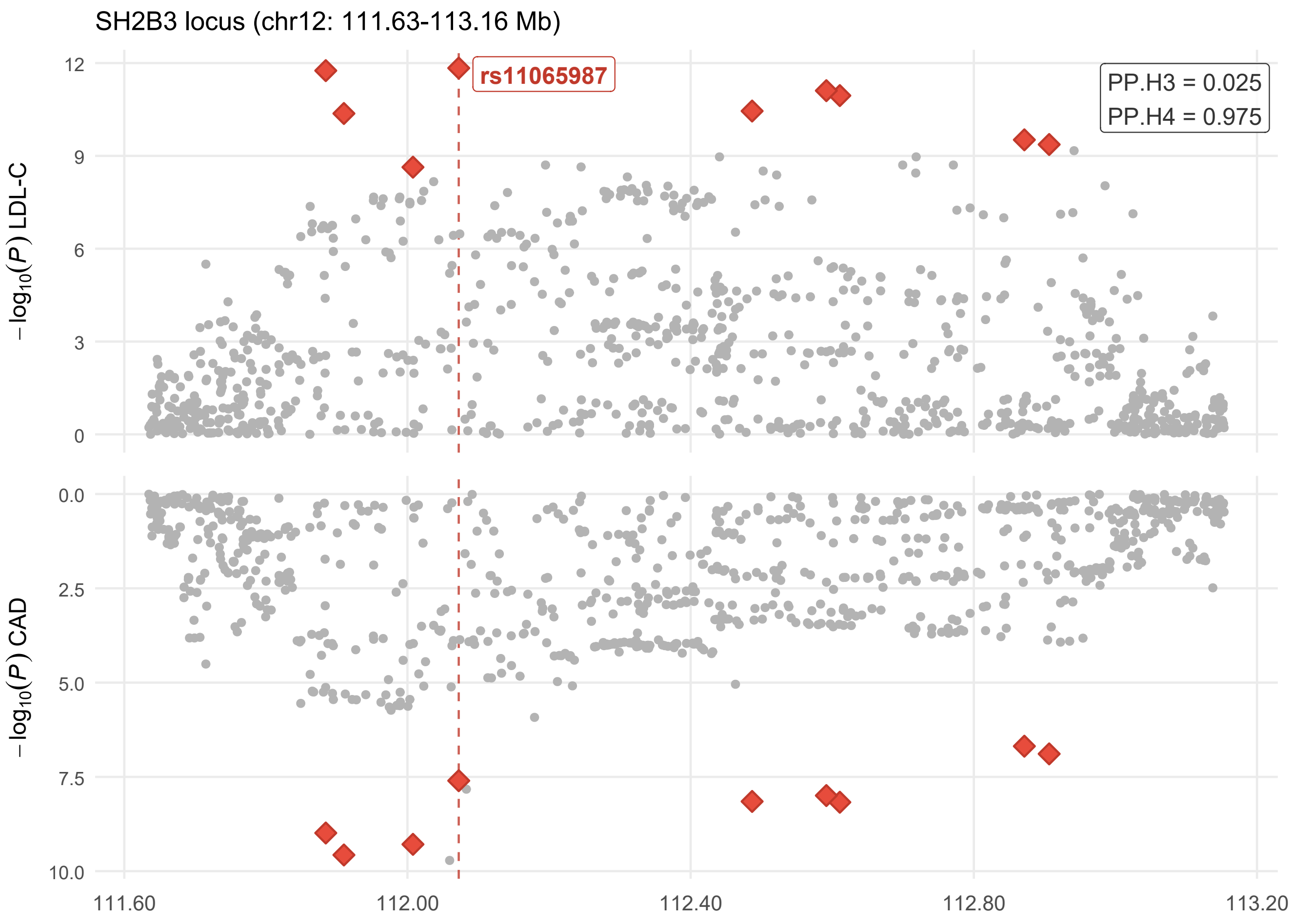

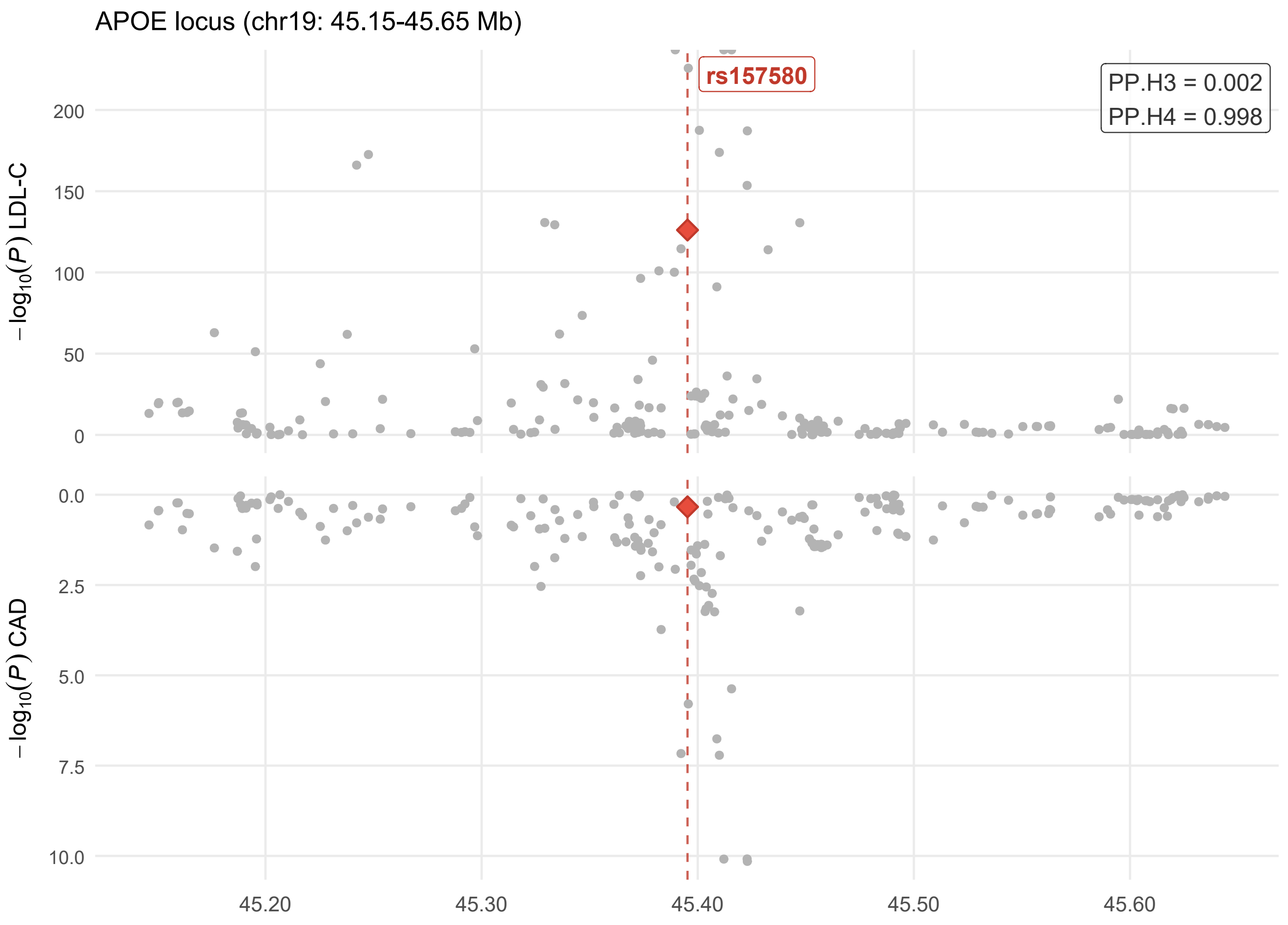

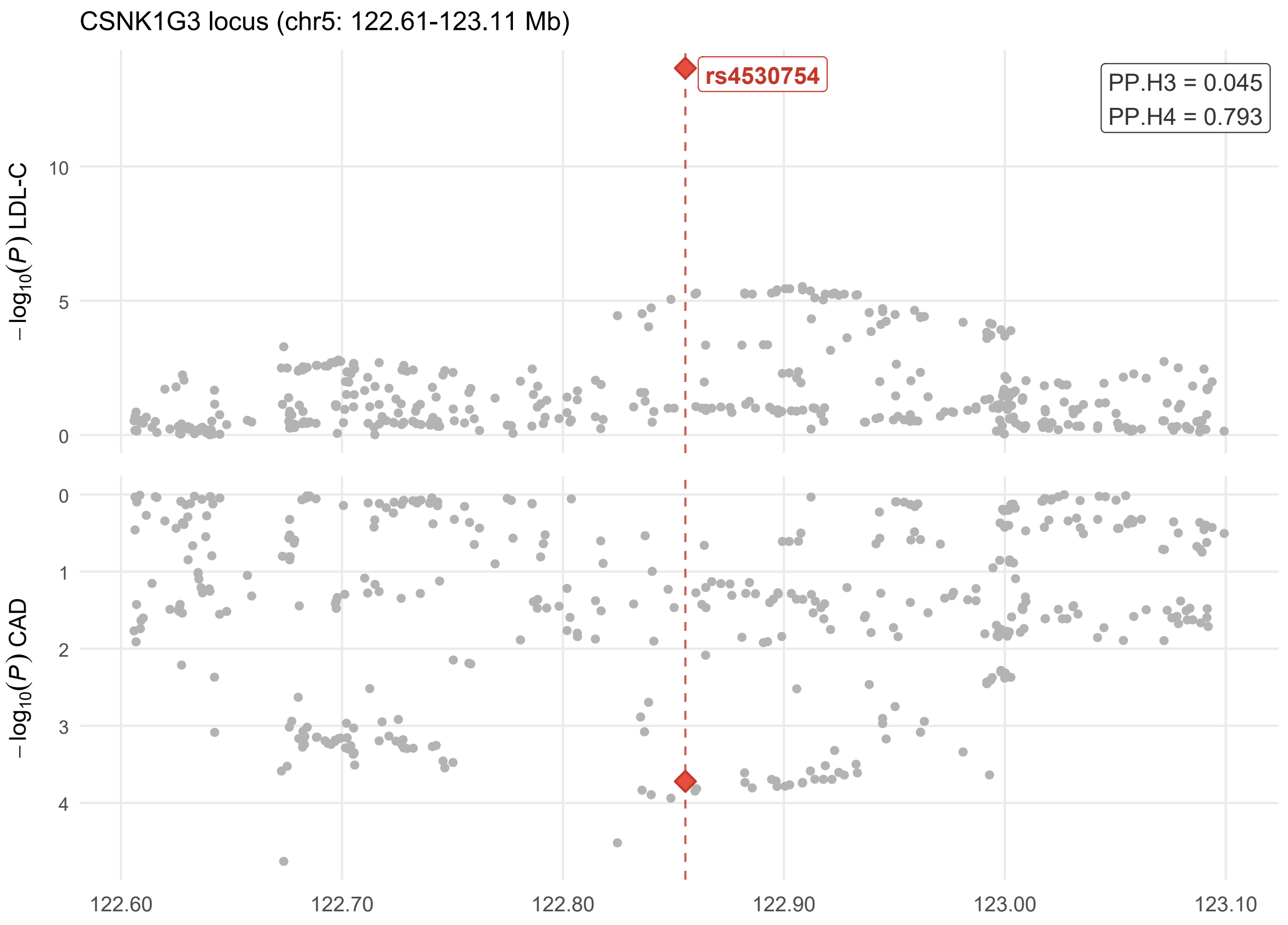

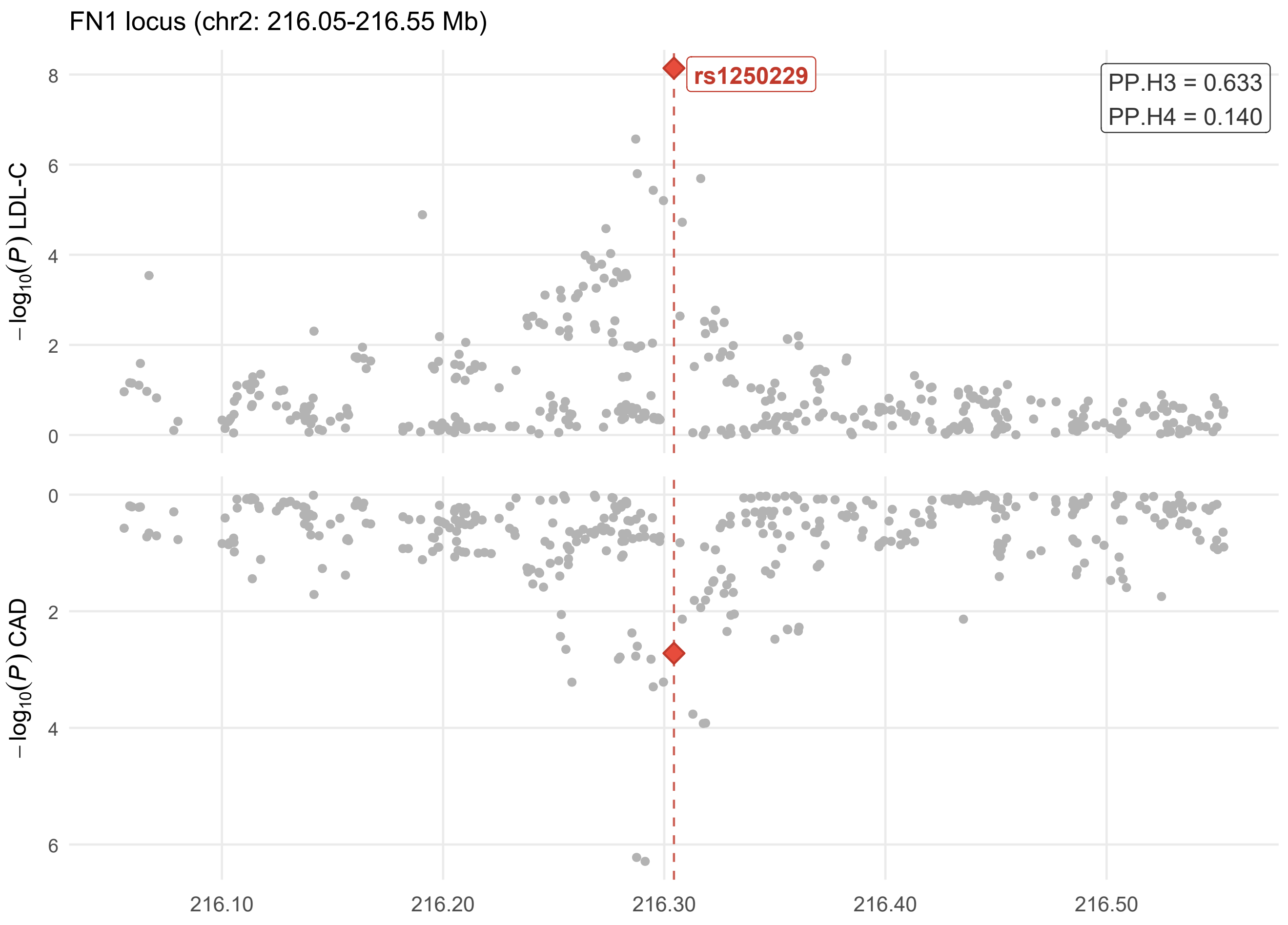
